# Direct haplotype-resolved 5-base HiFi sequencing for genome-wide profiling of hypermethylation outliers in a rare disease cohort

**DOI:** 10.1101/2022.09.12.22279739

**Authors:** Warren A Cheung, Adam F Johnson, William J Rowell, Emily Farrow, Richard Hall, Ana SA Cohen, John C Means, Tricia Zion, Daniel M Portik, Christopher T Saunders, Boryana Koseva, Chengpeng Bi, Tina Truong, Carl Schwendinger-Schreck, Byunggil Yoo, Jeffrey J Johnston, Margaret Gibson, Gilad Evrony, William B Rizzo, Isabelle Thiffault, Scott T Younger, Tom Curran, Aaron M Wenger, Elin Grundberg, Tomi Pastinen

## Abstract

Long-read HiFi genome sequencing (GS) allows for accurate detection and direct phasing of single nucleotide variants (SNV), indels, and structural variants (SV). Recent algorithmic development enables simultaneous detection of CpG methylation (mCpG) for analysis of regulatory element (RE) activity directly in HiFi-GS. We generated a comprehensive haplotype-resolved HiFi-GS dataset from a rare disease cohort of 276 samples in 152 families to identify rare (∼0.5%) hyper-mCpG events. We found that 80% of these events are allele-specific and predicted to cause loss of RE (LRE). We demonstrated heritability of extreme hyper-mCpG including rare *cis* SNVs and SVs causing short (∼200bp) and large hyper-mCpG events (>1 kb), respectively. We identified novel repeat expansions in proximal promoters predicting allelic gene silencing via hyper-mCpG and demonstrated allelic transcriptional events downstream. On average 30-40 LREs overlapped rare disease genes per patient, providing indications for variation prioritization. LRE led to a previously undiagnosed pathogenic allele in *DIP2B* causing global developmental delay. We propose that use of HiFi-GS in unsolved rare disease cases will allow detection of unconventional diseases alleles due to LRE.

## Introduction

Short-read exome (srES) or genome (srGS) sequencing is the tool of choice for the detection of single nucleotide variants (SNVs) in most human genetic applications. However, even with strict case selection for rare genetic disease in a clinical trial, srGS achieved only a ∼30% diagnostic rate ^1^, leaving most rare disease cases unsolved. On the other hand, 3^rd^ generation long-read platforms, such as PacBio’s Single Molecule, Real-Time (SMRT) HiFi-GS technology generating 12 – 16 kb reads, have been demonstrated to produce not only high quality SNV calls in difficult-to-map regions but also to accurately detect structural variants (SVs) genome-wide ^2^.

Within our large pediatric rare disease program, Genomic Answers for Kids (GA4K), collecting genomic data and health information for families with a suspected genetic disorder, we have integrated an enhanced sequencing pipeline using HiFi-GS into the routine follow-up of unsolved cases. Our pilot phase of ∼1000 families revealed that incorporating SVs from HiFi-GS resulted in new diagnoses in up to 13% of previously unsolved cases and HiFi-GS increased discovery rate of rare coding SVs >4-fold compared with srGS ^3^. However, the relative impact on undiagnosed rare disease due to incomplete interpretation of detected variants such as those mapping to non-coding regions remains unknown.

We have shown that non-coding SNVs can have pervasive effect on genome function and regulatory element (RE) activity, from splicing variation ^4,5^ to transcript levels in tissues and primary cells, ^6,7^ as well as chromatin-states ^8^ and CpG methylation (mCpG) levels ^9,10^. We exploited heterozygosity of functional alleles to augment the detection of differential RE activity ^11^ in the case of rare disease and for common population SNVs. We have also quantified other allele silencing effects, such as imprinting or X-inactivation that are measurable in chromatin, mCpG and gene expression data ^12-14^. We also previously showed that allelic RE hyper-mCpG reflects allelic regulatory/gene silencing and that genetic effects altering mCpG are more likely to be shared across tissues ^15,16^.

Established hyper-mCpG signatures are linked to several monogenic diseases ^17,18^ including imprinting disorders ^19^ and disorders caused by defects in chromatin regulators ^20^. In addition, “epimutations” have emerged through locus specific investigations, where a subset of missing rare disease alleles were shown to be non-coding and lead to hyper-mCpG and promoter inactivation ^21^. In most known cases of disease-impacting hyper-mCpG the effect is restricted to one allele, but even higher resolution techniques using short-reads for mCpG (whole-genome bisulfite sequencing, WGBS) ^16^ lack resolution for genome-wide analysis of allele-specific effects.

A recent HiFi-GS production pipeline update (Sequel IIe system release v11, PacBio, Menlo Park, CA) combines a deep learning model that integrates sequencing kinetics and base context, to generate mCpG profiles genome-wide from standard sequencing libraries. The augmented 5-base HiFi-GS platform allows single-molecular resolution of mCpG together with phasing from long contiguous accurate reads, with the potential to detect allele-specific events at an order of magnitude increased efficiency as compared to WGBS.

In this work we leverage the large GA4K rare disease program and generate a comprehensive HiFi-GS data set from 276 enrolled participants for parallel variant and mCpG calling. We validate the method through comparisons with WGBS and show high concordance in mCpG. We use the HiFi-GS data set to identify >50,000 rare hyper-mCpG event that are to a large extent (80%) allele-specific and predicted to cause loss of RE (LRE). We exemplify the power of HiFi-GS in unsolved rare disease cases by identifying a hyper-mCpG event that caused LRE and led to a previously undiagnosed pathogenic allele in *DIP2B* causing global developmental delay.

## Results

### Benchmarking HiFi-GS for parallel genome and methylome assessments in a rare disease cohort

We used our large GA4K cohort^3^ and generated a comprehensive mCpG dataset from 1367 enrolled participants utilizing WGBS (N=1184) and HiFi-GS (N=276) in peripheral whole-blood samples (**Supplementary Table 1**). Specifically, 1091 participants were profiled by WGBS only, 183 by HiFi-GS only and 93 by both platforms, respectively (**Supplementary Table 2)**. As a first validation step, we assessed the effect of technical variability on mCpG profiles by comparing the 93 samples profiled by both WGBS and HiFi-GS. We extracted 16.9 million paired CpGs sites (>20x coverage) for sample-wise correlations and found remarkable consistency across the two methods (median Pearson R=0.90, **Supplementary Table 3**). In addition, we evaluated the accuracy of mCpG profiling by extracting the top 500 most variable autosomal CpGs based on the HiFi-GS samples (**Supplementary Table 2**) for CpG-based correlations. Similarly, we noted high correlations compared to randomly permuted values (median Pearson R=0.86; **Supplementary Fig. 1; Methods**).

Next, we used individuals with unstable repeat disorders included in our data set as positive controls to test how genome-wide mCpG data by WGBS performed at known disease relevant regions. Specifically, we selected enrolled participants with *FXN* (**Fig. 1a**) and *FMR1* (**Fig. 1b**) mutations causing Friedreich ataxia (FRDA1)^23^ and Fragile X syndrome^22^, respectively, including two newly identified *FXN* intron 1 expansion carriers (**Fig. 1a**). While WGBS accurately identified hyper-mCpGs footprints in all carriers, the approach was not capable of reading into the pathogenic repeat or resolve carrier mCpG from homozygous sample. To contrast, we then applied HiFi-GS on additional probands with unstable repeat disorders including a case with a *DMPK* repeat expansion causing congenital Myotonic dystrophy type 1 (DM1)^18^. Here, haplotype-resolved HiFi-GS coupled with simultaneous high resolution mCpG profiling allowed us to not only detect the *DMPK* repeat expansion but also its associated large (∼1kb) hyper-mCpG signature (**Fig. 2**).

**Figure 1.**
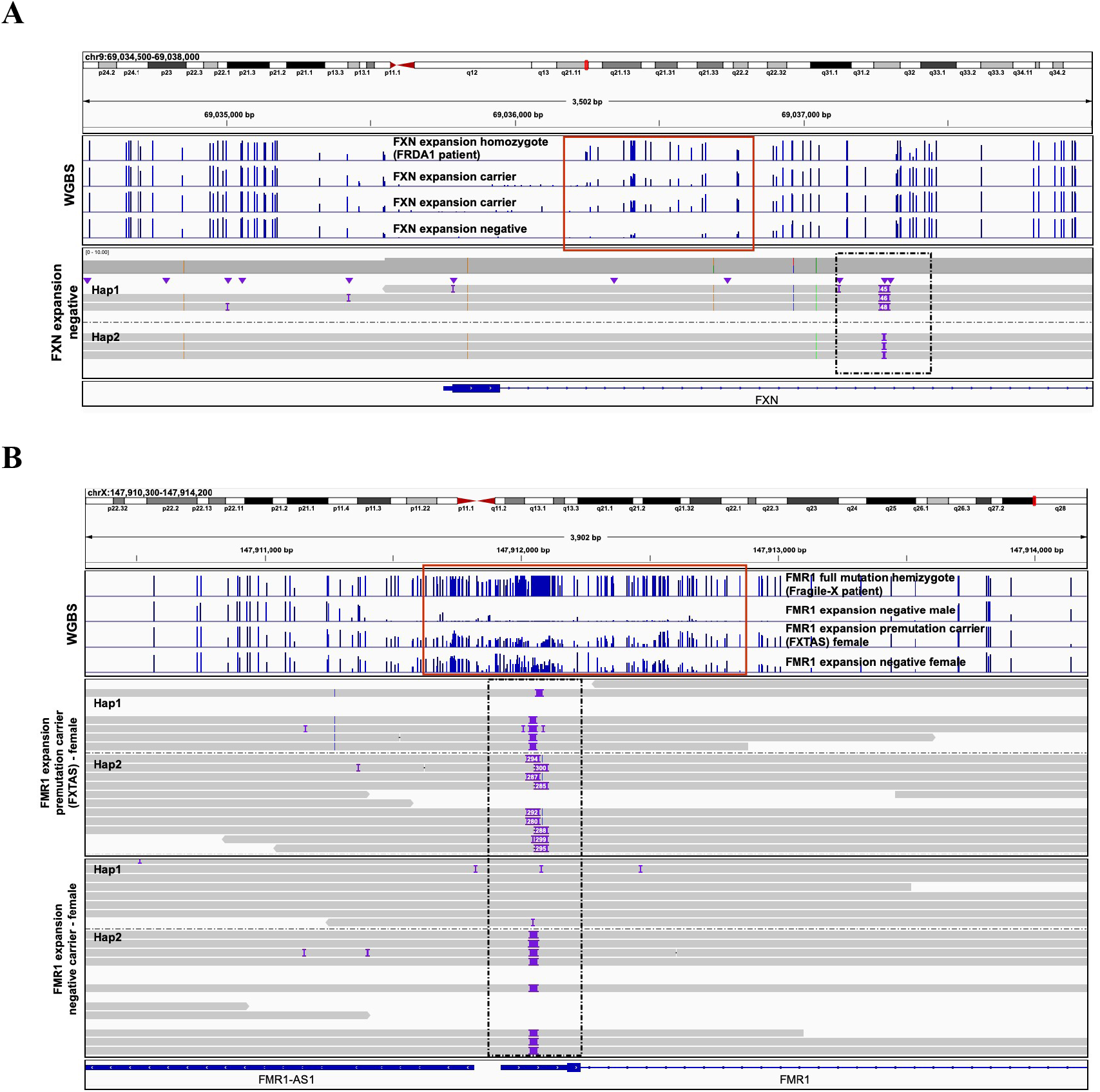
Whole-genome bisulfite sequencing (WGBS) and HiFi-Genome Sequencing (HiFi-GS) in patients with unstable repeat disorders. **A**. Genomic view of 2.7kb on chromosome 9 at the *FXN* locus showing one individual homozygous for the *FXN* expansion at pathogenic range (>65 repeats) causing the autosomal recessive disorder Friedrich’s Ataxia (FRDA1), two *FXN* expansion carriers and one individual without the expansion at a pathogenic range (<65 repeats, *FXN* expansion negative). Blue tracks show methylation level of CpGs measured by WGBS (y-axis, 0-100%) with hypermethylation footprint (red box) linked to pathogenic repeat expansion (>65 repeats). Grey tracks show haplotype-resolved repeat expansion by HiFi-GS (black box, dashed line) in an individual without the expansion at a pathogenic range (<65 repeats, *FXN* expansion negative). **B**. Genomic view of 3.7kb on X chromosome at the *FMR1* locus showing an hemizygote individual with Fragile X (>200 repeats), one female *FMR1* premutation carrier (55-200 repeats) and two individuals without the *FMR1* expansion (male and female, <55 repeats). Blue tracks show methylation level of CpGs measured by WGBS (y-axis, 0-100%) with hypermethylation footprint (red box) linked to pathogenic repeat expansion (>200 repeats). Grey tracks show haplotype-resolved *FMR1* repeat expansion (black box, dashed line) by HiFi-GS in one premutation carrier and one expansion negative female. Hap 1 denotes haplotype 1 and Hap 2 denotes haplotype 2.

**Figure 2.**
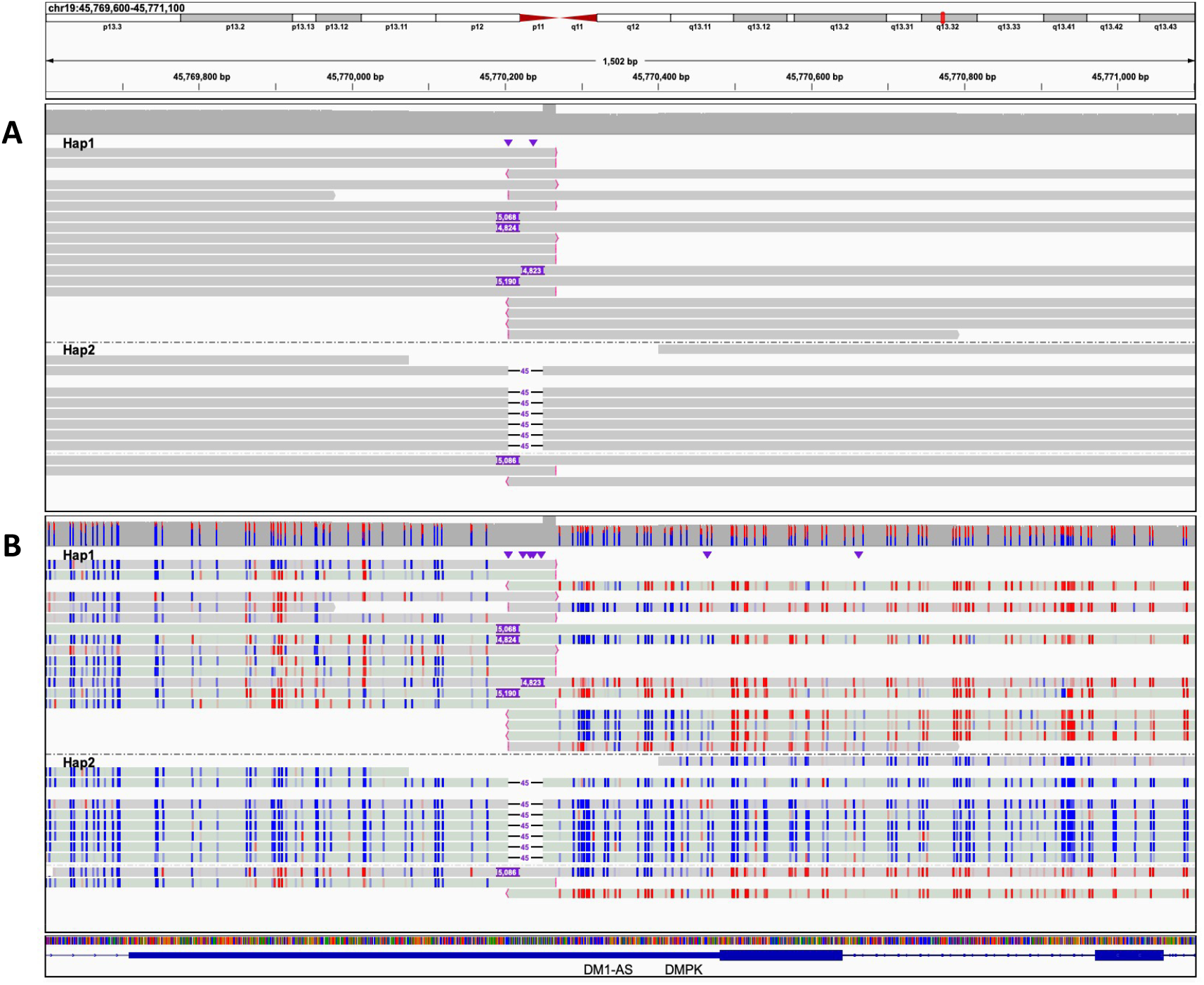
HiFi-Genome Sequencing (GS) in a Myotonic dystrophy type 1 (DM1) patient with *DMPK* repeat. Genomic view of 1.5kb on chromosome 19 at the *DMPK* locus showing an individual with the autosomal dominant disorder DM1. **A**. Haplotype-resolved raw HiFi-GS reads identifying “soft-clipped” reads (i.e., reads with red ends indicative of sequence not matching the reference) and variable sized insertion (4,823bp-5,190) on haplotype 1 (Hap 2) and a 45bp deletion on haplotype 2 (Hap 2). **B**. Haplotype-resolved HiFi-GS reads with CpG modification staining (blue indicating low CpG methylation prediction and red indicating high CpG methylation prediction). Soft-clipped and insertion (expansion) spanning reads show consistent hypermethylation flanking the disease allele.

Finally, we further demonstrated that haplotype-resolved mCpG across differentially methylated regions (DMRs) can be broadly utilized for the diagnosis of imprinting disorders ^24^. We exemplified this resolution at causal regions linked to Albright hereditary osteodystrophy ^25^ (*GNAS-AS1*; **Supplementary Fig. 2**), transient neonatal diabetes mellitus ^26^ (*PLAGL1*; **Supplementary Fig. 3**), Schaaf-Yang syndrome ^27^ (*MAGEL2;* **Supplementary Fig**. 4) and Temple Syndrome ^28^ (*MEG3/DLK1*; **Supplementary Fig. 5**).

### Identification and characterization of rare hypermethylation outlier event in rare disease cases

As shown for unstable repeat disorders, hyper-CpG can induce transcriptional silencing of disease genes via inactivation or loss of RE (LRE). Thus, we hypothesized that screening for outlying hyper-mCpG events genome-wide may aid the identification of coding and non-coding, functional rare SNVs and SVs in unsolved rare disease cases to ultimately improve diagnosis rate. We defined extreme hyper-mCpG outliers in two ways (**Fig. 3**). First, we ranked each CpG per individual based on the mCpG distribution for the population to catalog extreme hyper-mCpGs (**Methods**) and then classified hyper-CpG tiles (200bp) if two or more hyper-CpGs in the tile were extreme. The hyper-CpG tile was further classified as rare if it was only present in two or less unrelated individuals. Rare hyper-CpG tiles were then filtered reporting only those where the average z-score of all the CpGs of the tile was two or more. Using these criteria, we found a total of 25,543 extreme hyper-mCpGs tiles (**Supplementary Table 4)** with on average 125 hyper-mCpGs tiles per individual (**Supplementary Table 5)**. Quantity of extreme hyper-mCpGs tiles per individual did not differ among unaffected and affected after taking sequencing coverage into account.

**Figure 3.**
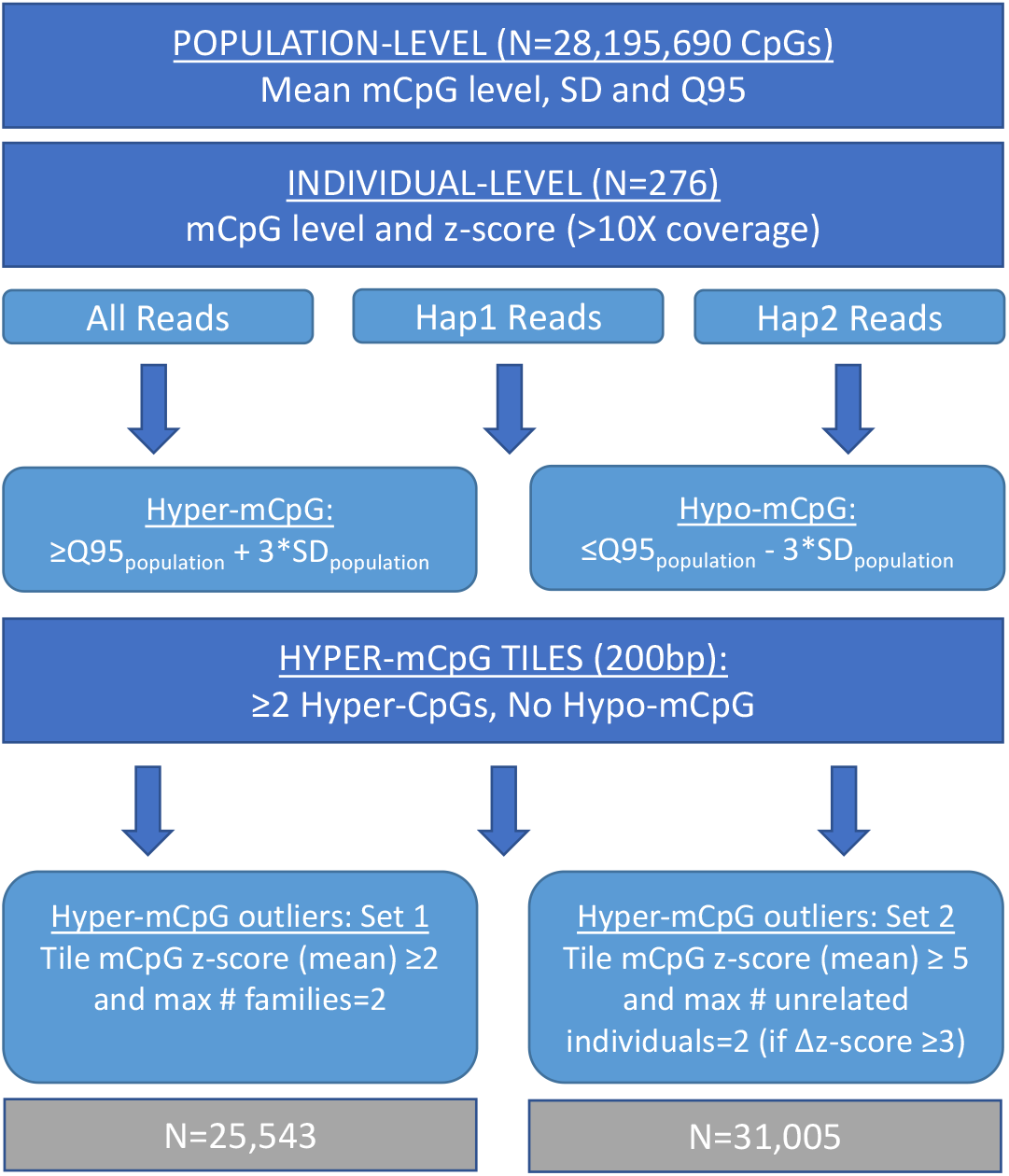
Simplified Schematic Depicting the Process to Identify Rare Hypermethylation Outliers in a Rare Disease Cohort.

However, these conservative measures of extreme hyper-mCpGs tiles reduce true discovery at dynamic regions where mCpGs show high degree of variation in the population (**Fig. 3**). To extract unusual observations among such dynamic mCpGs tiles, we allowed hyper-mCpGs tiles to be present in three or more unrelated individuals (removed the rarity criteria), but only kept instances where the average z-score of the CpGs in the hyper-mCpG tile was five or greater and with a minimum separation of three z-score units across unrelated samples allowing only a maximum of two samples per tile. These additional criteria retained another 31,005 hyper-mCpGs unique tiles (**Supplementary Table 6)** and on average an additional 113 extreme hyper-mCpGs per individual (**Supplementary Table 7)**.

Using the combined set of 56,548 unique hyper-mCpG tiles, we focused our analyses on the HiFi-GS subset carried out in affected patients (173 patients from 152 families**; Supplementary Table 1**) to provide the first genome-wide characterization of rare hyper-mCpG events in a rare disease cohort. This restricted the set of rare hyper-mCpG tiles to 30,672 hyper-mCpGs (**Supplementary Table 8**). We assessed the haploid dependency of the extreme hyper-mCpGs (i.e., the outlier effect being observed on one allele only) which yielded 80% allele-specificity (**Supplementary Table 8)**.

To further assess potential genetic underpinnings of rare hyper-mCpG tiles we estimated the degree of sharing of same hyper-mCpGs tile among related patients and observed a 3.1-fold enrichment (p<0.05; **Methods**). Next, we queried the closest SNV (minor allele frequency, MAF, <0.5%) and SVs (MAF<1%) for each hyper-mCpG tile and repeated the same test for all other mCpG tiles as a comparison. Using an empirical q value of less than 10% (**Methods**), we found that causal rare SNVs and rare SVs linked to the hyper-mCpG tiles were located within ∼1-2kb and within 10kb for SNVs and SVs, respectively (**Fig. 4a-b**). To bolster the potential causality for local rare genetic variation we exploited the haploid subset of hyper-mCpG tiles coupled with phased rare SNVs from the same reads (**Fig. 4c**) and showed statistically significant enrichment for local rare *cis-*SNVs extending up to 1kb (**Fig. 4d**) from the hyper-CpG tile. However, we noted strongest enrichment for SNV mapping close (<200bp) to the hyper-mCpG bin (**Fig. 4e**). Although WGBS were not able to resolve SNVs, it accurately detected associated hyper-mCpG footprints in probands carrying the rare SNVs (**Supplementary Fig. 6-8**). However, for local SVs and insertion (INS)-deletions (DELs), variations in reads are not phased (**Methods**) and required manual curation. For 40 randomly chosen unphased short DELs called by DeepVariant and verified in read data and mapping near hyper-mCpG tiles, we observed 31 out of the 40 (binomial P= 0.0007) in *cis* suggesting similar behavior as for SNVs with respect to allelic and distance distribution (**Supplementary Fig. 9**).

**Figure 4.**
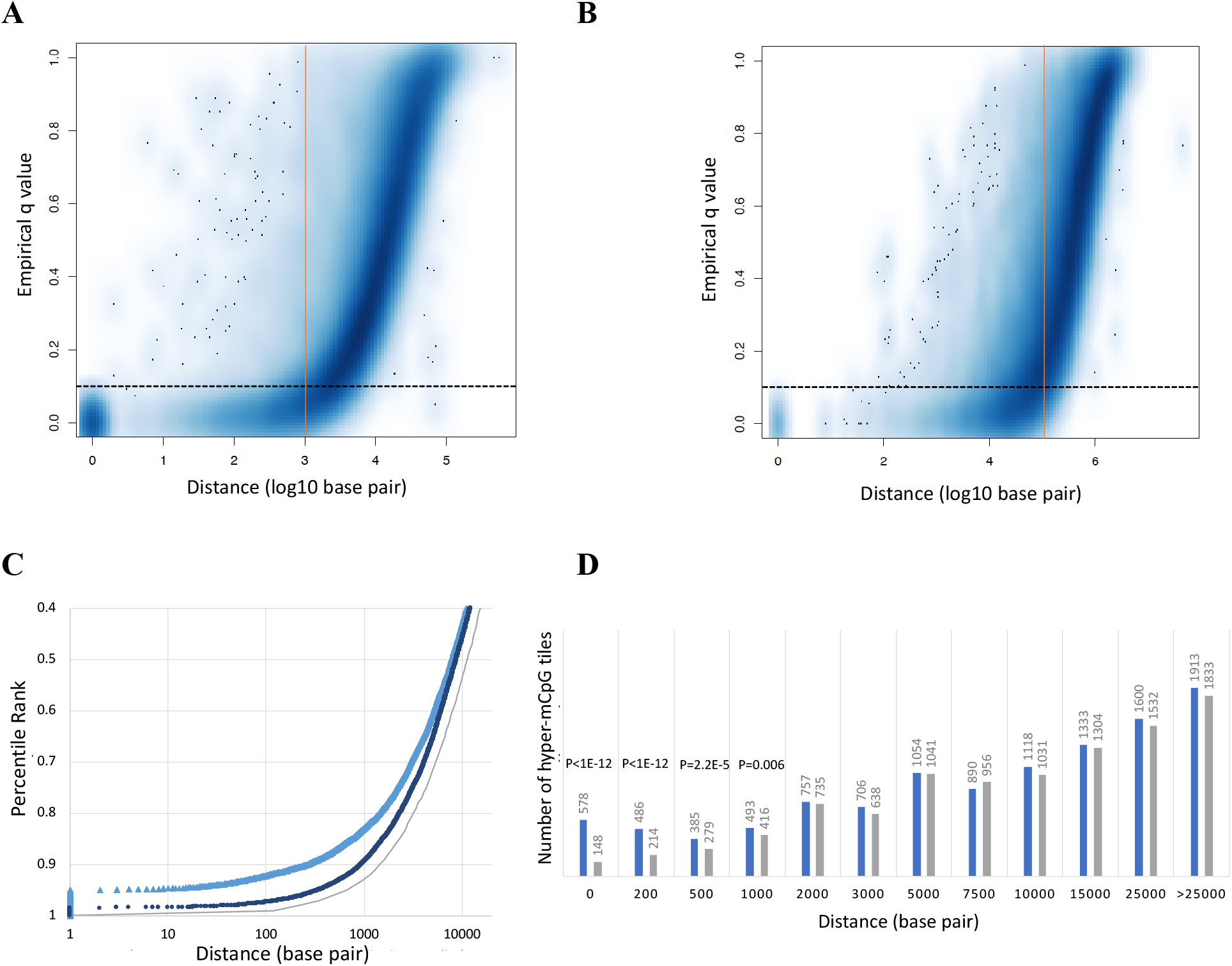
Genetic Causes of Rare Hypermethylation Outliers. Scatter plot depicting the distance (x-axis, log10 base pair(bp)) between rare hyper-mCpG tiles and closest rare **A**. SNV (<0.5% MAF gnomAD) and **B**. SVs (<1% MAF in HiFi-GS population data). Same process was repeated for, in parallel, for same tile in all other individuals to obtain a percentile rank of tile-variant distances in the population. Empirical q value (y-axis) threshold (dashed black line, q<0.1) corresponding to estimated distance of causal rare variant of rare hyper-mCpG tiles (red solid line). **C**. Scatter plot depicting haplotype-resolved distance (x-axis, bp) between rare hyper-mCpG tiles and closest rare SNV (0.5% MAF gnomAD) in *cis* (same read, light blue triangle) and *trans* (opposite read, dark blue round). Same process was repeated for, in parallel, for same tile in all other individuals to obtain a percentile rank (y-axis) of tile-variant distances in the population. Grey line depicts null distance distribution of rare SNVs (<0.5% MAF gnomAD) in our sample set. **D**. Bar graph depicting number of extreme hyper-mCpG tiles (y-axis) linked to rare SNV in *cis* (same read, blue bars) or *trans* (opposite read, grey bars) based on distance. P-values from binomial test assuming equal number of tiles in *cis* and *trans* for at each distance (x-axis, bp). **E**. Genomic view of an example of a rare SNV (black box) mapping in *cis* close to a hyper-mCpG tile on chromosome 10 causing allele-specific hypermethylation. Track depicts haplotype-resolved HiFi-GS reads with CpG modification staining (blue indicating low methylation prediction and red indicating high methylation prediction). Hap 1 denotes haplotype 1 and Hap 2 denotes haplotype 2.

Finally, most outlier effects were restricted to a few hundred bp corresponding to two or less hyper-mCpG tiles referred to as ‘regular’ hyper-mCpG tiles (**Supplementary Fig. 10**). However, 4% of the hyper-mCpG tiles were linked to larger mCpG perturbation (i.e., two or more hyper-mCpG tiles referred to as ‘large’ hyper-mCpG tiles) and among such extended signals, local SV or SNV (<1kb) were significantly more common than expected based on their overall distribution. Specifically, we found that 129 out of 783 (16%) ‘large’ hyper-mCpG tiles had a local SVs or SNVs mapped within 1kb whereas only 96 out of 1057 (9%) ‘regular’ hyper-mCpG tiles had a local SVs or SNVs mapped within the same distance (i.e., 1kb).

### Rare genetic variation links to novel hypermethylation in regulatory elements

To assess the extent to which the identified hyper-mCpGs outlier tiles (N=56,548) map to RE we used the largest atlas of regulatory DNA to date that was created from 16 different human tissue types or states^35^. We found that 94% of the extreme hyper-mCpG outliers mapped to a RE compared to 41% for randomly selected non-outlier mCpG tiles (**Methods**), representing a ∼2-fold enrichment (**Fig. 5**). We noted largest enrichment for RE 1) specific to immune cells (myeloid, erythroid, and lymphoid) in line with the tissue type used for HiFi-GS and 2) shared across 10 or more organ systems (tissue invariant) (**Fig. 5a**). As expected, given the high degree of allele-specificity as discussed above, many of these hyper-mCpGs outliers overlapping a RE are linked to rare genetic variation in *cis* mapping either proximal or distal to their potential cognate target genes. For instance, we identified a rare 2.6kb insertion (INS) (**Fig. 5b)** in *cis* causing proximal RE hypermethylation at the *DDB2* disease locus (**Fig. 5c**). Similar effects on RE were observed for SNVs (**Supplementary Fig. 11**) and SVs (**Supplementary Fig. 12**) as well as for novel repeat expansions (**Supplementary Fig. 13-15**), duplications (**Supplementary Fig. 16)** and deletions (DELs) (**Supplementary Fig. 17-18)**.

**Figure 5.**
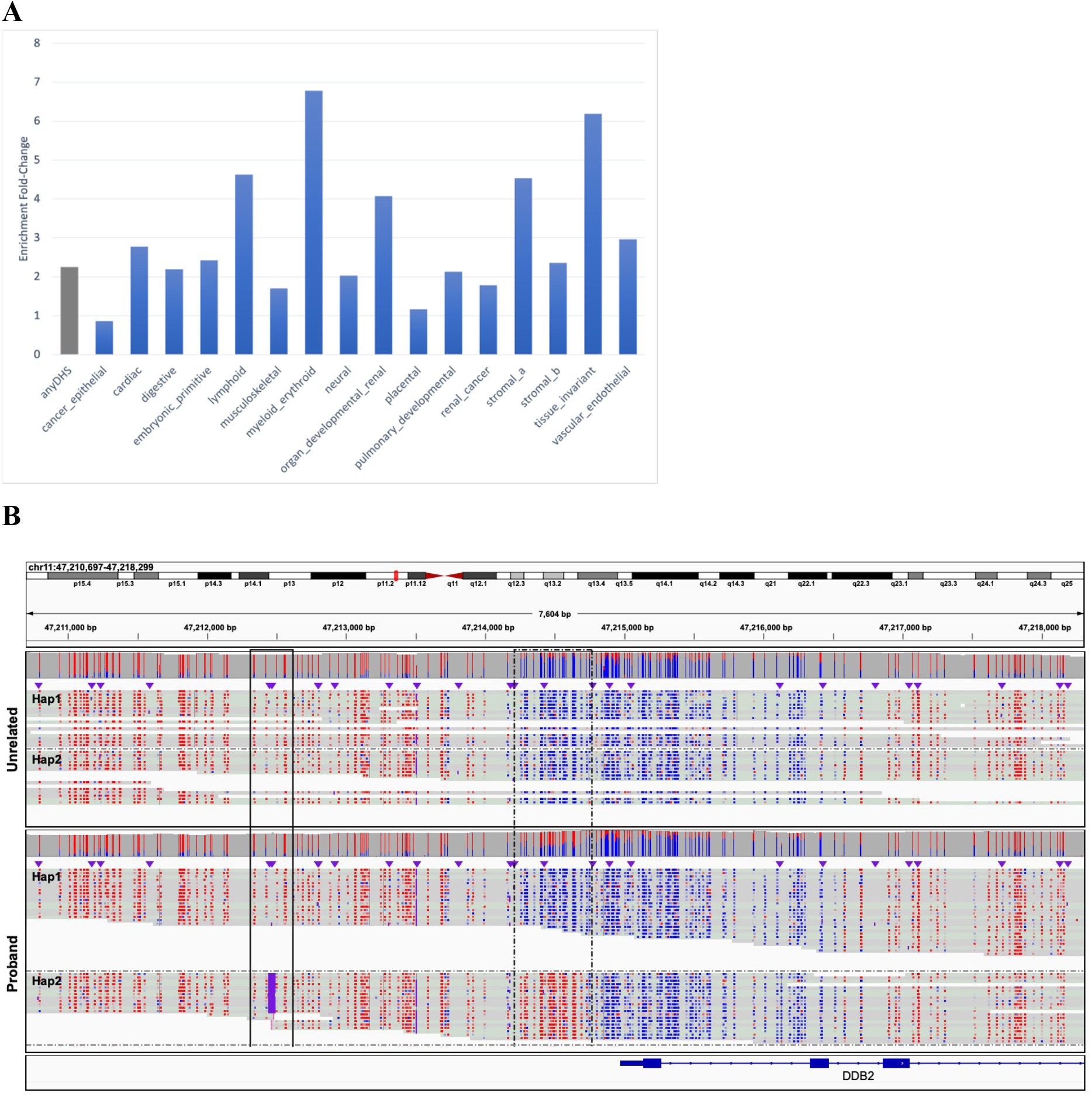
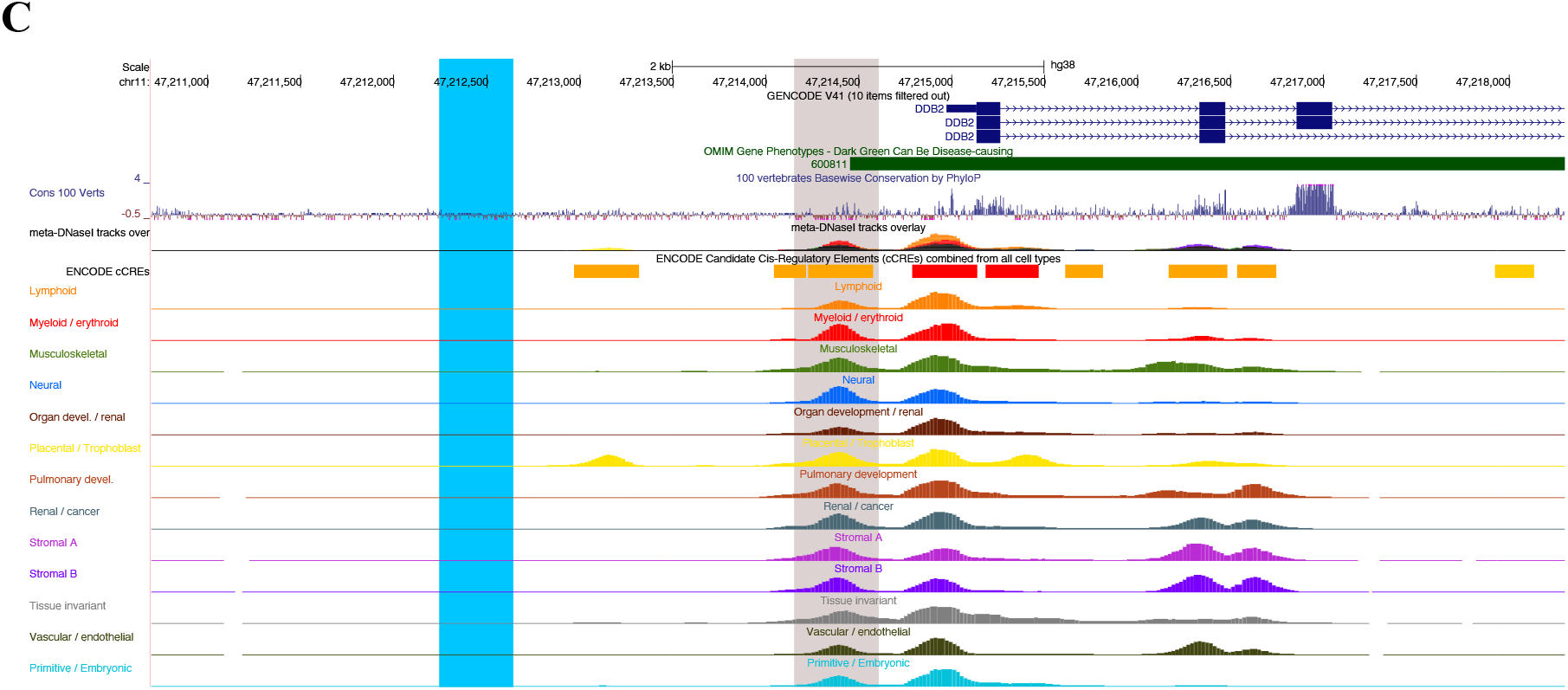
Regulatory Element Annotation of Rare Hypermethylation Outliers. **A**. Bar graph depicting the fold-enrichment (y-axis) of the overlap of regulatory elements (assessed by DNase I hypersensitive site (DHS) mapping) with extreme hyper-mCpG tiles compared to background control tiles. Enrichment is presented as either overlapping any DHS site (grey bar) or specific to tissue states (blue bars). **B**. Genomics view of 7.6kb at the *DDB2* disease locus comprising an insertion (black box, solid like) resolved by HiFi-GS that results in 1kb proximal promoter hypermethylation (black box, dashed line). Haplotype-resolved CpG modification staining (blue indicating low methylation prediction and red indicating high methylation prediction) is shown in proband and unrelated sample. Hap1 denotes haplotype 1 and Hap2 denotes haplotype 2. **C**. Zoomed in genomics view depicting regulatory element (DHS) annotation of the hyper-mCpG tile (grey box) nearby causal insertion (blue box).

### Validation of rare hypermethylation events by isoform profiling

Since we focused on rare hyper-mCpG, it is anticipated that the large magnitude of outliers can predominantly be found in the relatively hypomethylated, dynamic ^29^ REs of the human genome. Consequently, these rare hyper-mCpG outlier may contribute to RE silencing and impact gene expression. To this end, we demonstrated the functional translation of mCpG in a subset of analyses from patients by co-occurrence of allelic rare hyper-mCpG events in proximal RE and concurrent allelic mRNA silencing. Specifically, we selected hyper-mCpGs outliers (**Supplementary Table 4**) that mapped within 1kb of a transcription start site (TSS) and present in an individual where long-read isoform sequencing (IsoSeq) was performed on proband-specific induced pluripotent stem cells (iPSCs). The TSS was linked to the closest consensus coding sequence (CCDS) region and IsoSeq reads informative for a heterozygous SNV were selected if covered at more than 10 reads for each selected proband and controls. The latter corresponded to all other probands not ‘carrying’ the hyper-mCpG tile. In total, 22 unique hyper-mCpG outliers-transcript pairs across eight probands were informative. We found that in total of 15/22 (68%) of the cases were associated with a differential abundance of the allelic copies of the tested transcript beyond a 45:55 ratio compared to cases where only 3/22 (14%) showed deviations beyond similar ratio (**Supplementary Table 9; Supplementary Fig. 19**). Notably, this was achieved in non-blood cells (iPSCs) from the same patients (**Methods**), indicating that some variation exhibits relative to tissue independence.

### Rare hypermethylation outlier facilitates diagnosis of unsolved rare disease cases

Having genetically and functionally characterized rare hyper-mCpGs tiles, we next queried their relationship with rare disease genes (OMIM) to explore candidate functional changes for unsolved disease. Among the patients, we observed a total of 5,438 hyper-mCpG tiles across 4,400 OMIM genes (∼30 per patient). We anticipate that these outliers can generate new non-coding disease variant candidates. Among the OMIM gene overlapping hyper-mCpG tiles, two of the top five highest z-scores mapped to *GNAO1* with a rare intronic SNV (C-to-A MAF<0.003) causing the hypermethylation footprint (∼500bp). The hyper-mCpG tiles map to a vertebrate constrained sequence and ENCODE predicted distal regulatory (enhancer) active across multiple tissue types (**Fig. 6a**). By accessing HiFi-GS from parents, we noted that both the proband and the father share the SNV with similar hypermethylation effect of the RE. *De novo* mutations in *GNAO1*, both gain of function and loss of function, cause neurodevelopmental disorder including developmental and epileptic encephalopathy. In this case, the proband (at 2 years) suffers from dysphagia and failure-to-thrive, with paternal family history of seizures. Considering the wide spectrum of *GNAO1* associated neurodevelopmental disease and the undiagnosed proband’s presentation and family history, the variant is a candidate for further study. However, with the expression pattern of *GNAO1* being almost exclusively brain specific (**Supplementary Fig. 20)**, follow-up analysis requires tissue-targeted experimentations in affected individuals.

**Figure 6.**
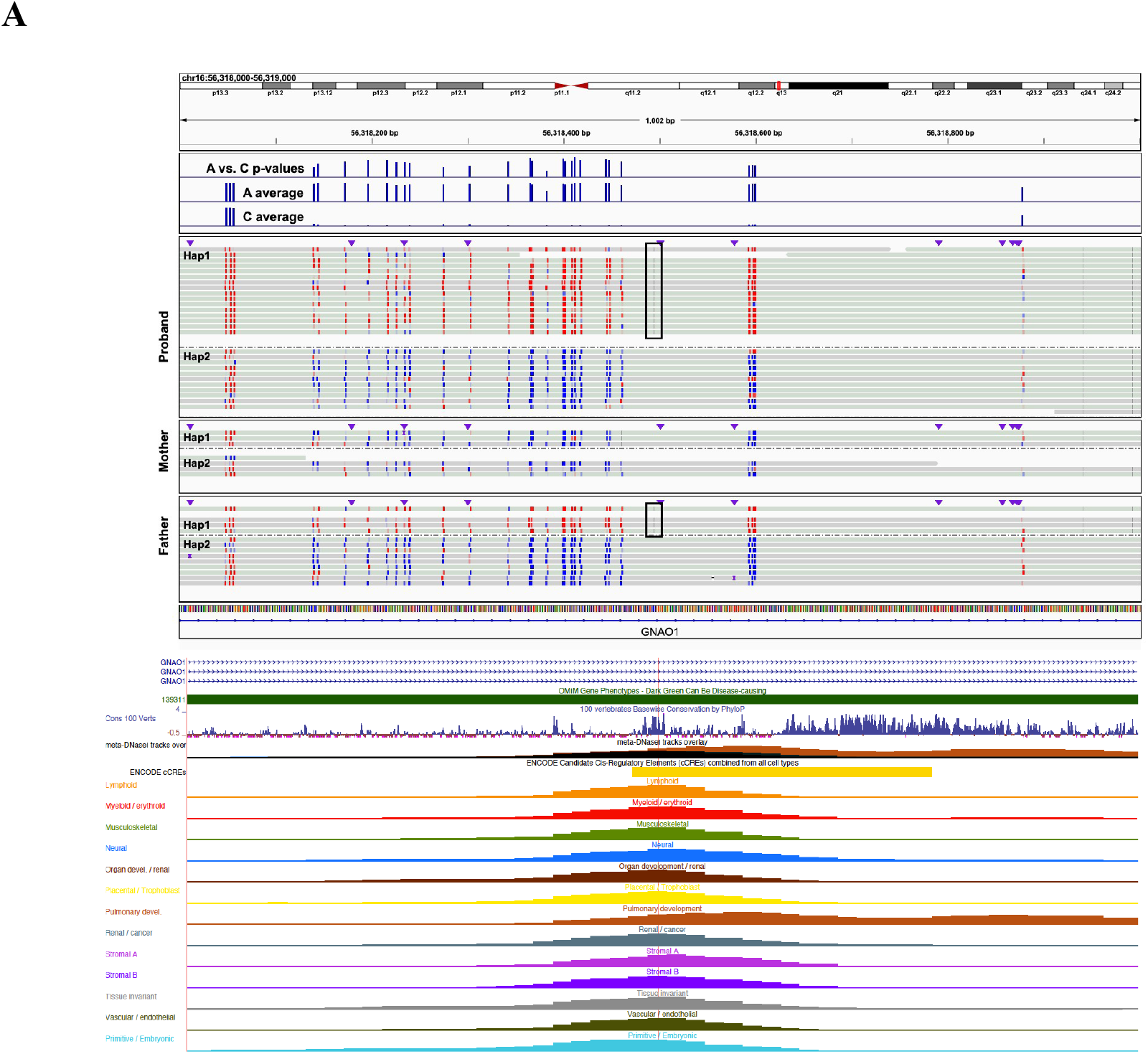
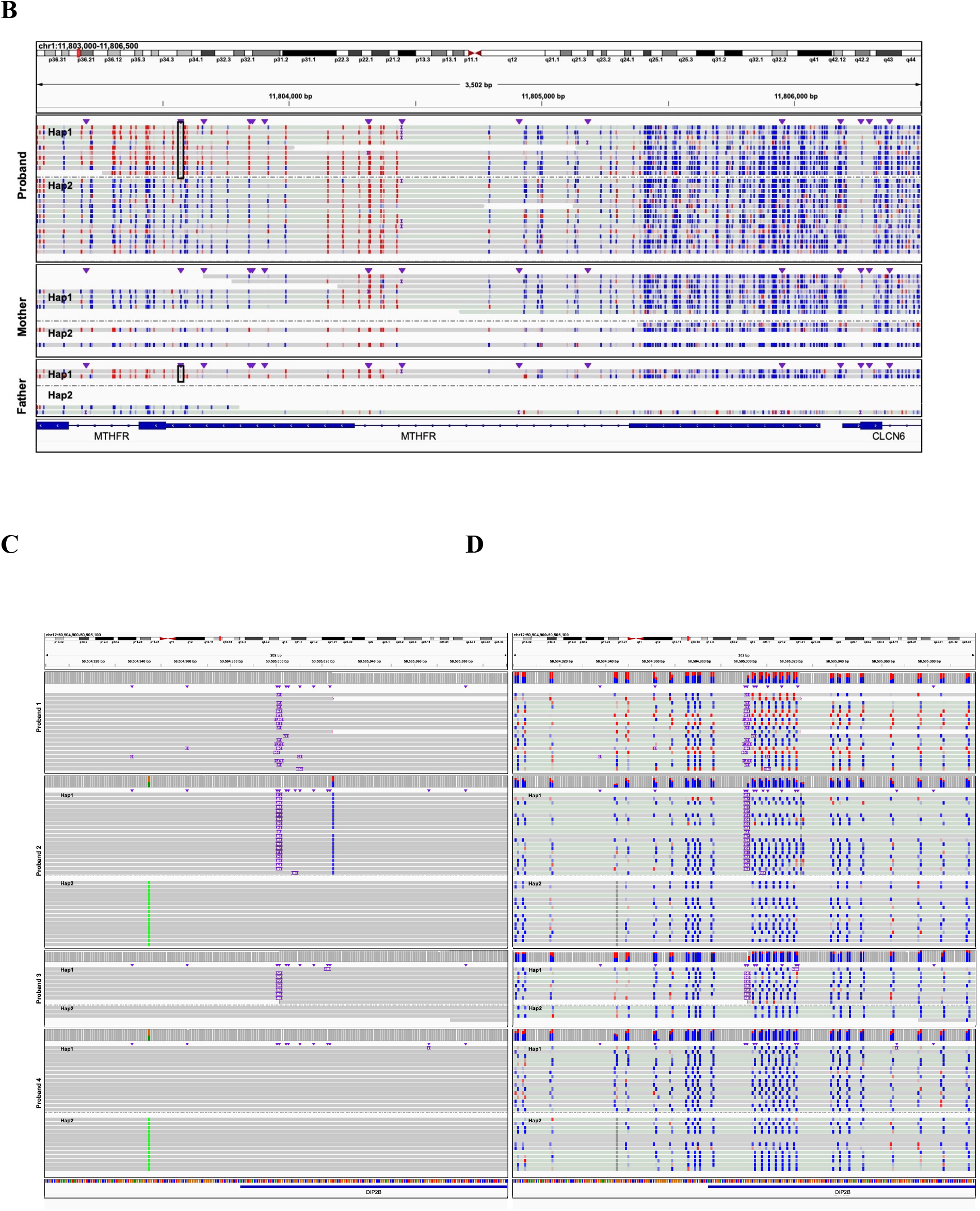
OMIM Genes with Rare Hypermethylation Event in Regulatory Elements. **A**. Genomic view of ∼1kb on chromosome 16 at the *GNAO1* locus showing a paternally inherited rare intronic SNV (C-to-A MAF<0.3%) at chr16:56,318,495 (black rectangle). Haplotype-resolved CpG modification staining (blue indicating low methylation prediction and red indicating high methylation prediction) is shown in the complete trio. Hap1 denotes haplotype 1 and Hap2 denotes haplotype 2 in respective sample. Top track shows P-values (y-axis, -log10(0-31)) from Fisher’s exact test for CpG methylation differences in rare A versus common C-allele carrying reads, respectively. Second and third track from the top shows average CpG methylation rate (y-axis, 0-100%) across all A (rare) and C (population prevalent) carrying reads, respectively. USCS genome browser view depicts the rare SNV (red line) overlapping a vertebrate constrained sequence and conserved regulatory element active across tissue types. **B**. Genomic view of ∼3.5kb on chromosome 1 at the *MTHFR* locus showing a paternally inherited rare 5’UTR SNV (black box) which maps into an extended ∼1kb extreme hyper-mCpG tile at the *MTHFR* proximal promoter. Haplotype-resolved CpG modification staining (blue indicating low methylation prediction and red indicating high methylation prediction) is shown in the complete trio. Hap1 denotes haplotype 1 and Hap2 denotes haplotype 2 in respective sample. **C**. HiFi-GS reads across four individuals detecting 1) large (Proband 1), 2 and 3) expanded (Haplotype-resolved, Proband 2 and Proband 3) and 4) no repeats (Haplotype-resolved, Proband 4) at the *DIP2B* locus. Hap1 denotes haplotype 1 and Hap2 denotes haplotype 2 in respective sample. **D**. CpG modification staining of HiFi-GS reads (blue indicating low methylation prediction and red indicating high methylation prediction) across the four individuals.

Next, we selected hyper-mCpG tiles close to the proximal promoters of OMIM genes (+/-1kb) which yielded 1,341 regions. We then further restricted to hyper-mCpG tiles with local *cis-* rare SNV or SV for a new subset of 66 regions and noted that most of these genetically linked hyper-mCpG events in our dataset occurred in autosomal recessive OMIM gene promoters. We exemplified this at the *MTHFR* locus showing a paternally inherited rare 5’UTR SNV associated with several hundred base pair of hyper-mCpG (**Fig. 2b**) and indicating carriership of a potentially deleterious allele. Another set of four hyper-mCpG events (in three patients) near OMIM genes were flagged for follow-up: one tile in *NSD1*, one in *SET* and two adjacent hyper-mCpGs tiles in *DIP2B*. Further analysis revealed that the hyper-mCpG of the proximal regulatory region of *DIP2B* was immediately adjacent to a previously missed and undiagnosed^3^ repeat expansion in a proband with global developmental delay (**Fig. 2c)**. Here, the phasing power and mCpG ‘staining’ ability by HiFi-GS (**Fig. 2d)** allowed for prioritization and further clinical review where the expansion was determined to be within the pathogenetic range (∼280-300 repeats, >250 considered pathogenic). This diagnostic finding was clinically validated by triplet repeat PCR (**Methods**) and reported to the provider. Within our large HiFi-GS cohort we had additional unsolved patient specimens with expanded *DIP2B* repeats (**Fig. 2d**), however all other expansions were below pathogenic range and remained hypo-mCpG – exemplifying the augmented power of long repeat resolving reads coupled with mCpG detection.

## Discussion

Rare genetic diseases remain a translational challenge in the era of advanced molecular diagnostics. Better data sharing and increasing understanding of expressivity through larger molecularly classified rare disease cohorts have the capacity to discover new genes and assign function to variants of unknown significance (VUS). However, structural genetic variation and non-coding variation with disease causing potential are areas of potential unrealized diagnostic yield due to technological and analytical hurdles. Suggestions for closing technology gaps include coupling next-generation sequencing (NGS) with additional assays gathering either transcriptomic ^30^ or epigenomic ^31^ data. Integrated genome-wide, high-resolution assessment of any two data modalities coupled with high haploid distinction has not been attempted to date. In our rare disease cohort, we demonstrate several key aspects of combined mCpG and HiFi-GS for exploration of disease variation. Known disease-linked and parent-of-origin mCpG is recovered alongside with full GS. Rare novel variation in the epigenome is heritable and can be shown to be physically linked to rare non-coding genetic variation in long reads. A substantial fraction of outlier signatures is caused by complex, local rare SV which is challenging to detect by NGS.

We demonstrate predicted silencing of REs represented by hyper-mCpG outliers propagating to transcriptional silencing in same patient but in distinct lineage cells, showing that blood-based 5-base HiFi-GS can have multi-tissue relevance. Employing an analytical approach to identifying population variation, we can limit the search to reasonable sets of candidate alleles for prioritization of manual curation where our stringent filter already identified previously missed diagnosis for a rare disease involving *DIP2B*. However, the potential discoveries by this platform, even within this dataset with follow-up validation, will expand to novel disease alleles and demonstrates a new ability to link non-coding variation to clinical evaluation in rare diseases.

## Online Methods

### Study Cohort

The study cohort described includes 1243 affected patients from 1078 families, with a total of 1367 individuals (detailed in **Supplemental Table 1**) enrolled in the Genomic Answers for Kids program ^3^. Patients with a suspected rare disease were referred from multiple different specialties, with the largest proportion nominated by Clinical Genetics, followed by Neurology. A continuum of pediatric conditions is represented, ranging from congenital anomalies to more subtle neurological and neurobehavioral clinical presentations later in childhood. Of the 1243 affected patients, 141 had a known genetic diagnosis at the initiation of the study.

### Sample collection and preparation

Whole blood was obtained from each study participant in EDTA and sodium heparin collection tubes for DNA and peripheral blood mononuclear cell isolation (PBMC), respectively. DNA was isolated using the chemagic™ 360 automated platform (PerkinElmer) and stored in -80°C. PBMCs were isolated using a RoboSep-S (StemCell) and the EasySep Direct Human PBMC Isolation Kit (StemCell 19654RF). After automated separation, the enriched cell fraction was centrifuged at 300 rcf for 8 minutes then resuspended in 1 mL of ACK Lysing Buffer (Thermo Fisher A1049201) and incubated at room temperature for 5 minutes. The cell suspension was diluted with 13 mL of PBS (Thermo Fisher 10010023) supplemented with 2% FBS (Cytiva SH30088.03HI) then centrifuged at 300 rcf for 8 minutes. The cell pellet was resuspended in 1 mL of PBS + 2% FBS, and cell count and viability were assessed using a Countess II automated cell counter (Thermo Fisher). Cells were centrifuged at 300 rcf for 8 minutes and resuspended in 1 mL of cold CryoStor CS10 (StemCell 07930) then transferred to a cryogenic storage vial. Cells were frozen slowly in a Corning CoolCell FTS30 placed at -80°C overnight then transferred to liquid nitrogen vapor the following day for long-term storage. Human patient-specific induced pluripotent stem cells (iPSCs) were generated from a subset of the patient’s PBMCs using episomal vectors. In brief, PBMCs were maintained in StemSpan SFEM II (STEMCELL Tech, 09605) media plus 1X Antibiotic-Antimycotic (ThermoFisher, 15240062) for 3-5 days, followed by nucleofection of the following episomal plasmids: pCXLE-hOCT3/4-shp53-F (Addgene, 27077), pCXLE-hSK (Addgene, 27078), pCXWB-EBNA1 (Addgene, 37624) and pCXLE-hUL (Addgene, 27080). After nucleofection, transfected PBMCs were plated onto 35-mm matrigel (Corning, 354277) coated tissue culture dishes in Stemspan SFEM II media supplemented with 10 μM Y-27632 (Tocris, 1254). Two days later, ReproTESR (STEMCELL Tech, 05926) was added and plates centrifuged at 50xg for 30 minutes. Every other day fresh ReproTESR was added until iPSC colonies were visible. Once colonies were visible ReLeSR (STEMCELL Tech, 05872) was used to isolate iPSC colonies. iPSC colonies were maintained in mTeSR1 complete media (STEMCELL Tech, 85850) on matrigel coated tissue culture plates and refed every other day or as needed until ready. Cells were cultured at 37 °C in 5% CO2.

### PacBio HiFi long-read genome sequencing and analysis

In total of ∼5 ug of DNA per sample was sheared to a target size of 14 kb using the Diagenode Megaruptor3 (Diagenode, Liege, Belgium). SMRTbell libraries were prepared with the SMRTbell Express Template Prep Kit 2.0 (100-938-900, Pacific Biosciences, Menlo Park, CA) following the manufacturer’s standard protocol (101-693-800) with some modifications as described previously ^3^. Libraries were sequenced on the Sequel IIe Systems using the Sequel II Binding Kit 2.0 (101-842-900) or 2.2 (102-089-000) and Sequel II Sequencing Kit 2.0 (101-820-200) with 30 hr movies/SMRT cell. Samples were sequenced to a target of >10X coverage. Circular consensus reads were generated with ccs v6.3 (https://github.com/PacificBiosciences/ccs) using the “--hifi-kinetics” option to generate consensus kinetics tags, and primrose v1.1 (https://github.com/PacificBiosciences/primrose) was used to predict 5mC modification of each CpG motif and generate base modification (“MM”) and base modification probability (“ML”) BAM tags. HiFi Read mapping, variant calling, and genome assembly were performed using a Snakemake workflow (https://github.com/PacificBiosciences/pb-human-wgs-workflow-snakemake). HiFi reads were mapped to GRCh38 (GCA_000001405.15) with pbmm2 v1.9 (https://github.com/PacificBiosciences/pbmm2). Structural variants were called with pbsv v2.8 (https://github.com/PacificBiosciences/pbsv) with “--hifi --tandem-repeats human_GRCh38_no_alt_analysis_set.trf.bed” options to pbsv discover and “--hifi -m 20” options to pbsv call. Small variants were called with DeepVariant v1.3 following DeepVariant best practices for PacBio reads (https://github.com/google/deepvariant/blob/r1.3/docs/deepvariant-pacbio-model-case-study.md) and locally phased with WhatsHap v1.0 ^32^. Local phase haplotype tags (“HP”) were added to the aligned BAM by WhatsHap v1.0. Pileup-based consensus methylation sites and probabilities were generated by the script “aligned_bam_to_cpg_scores.py” from pb-CpG-tools v1.1.0 (https://github.com/PacificBiosciences/pb-CpG-tools/) with the “-q 1 -m denovo -p model -c 10” options. Reads were visulaized in IGV version 2.15.2.

### Whole-genome bisulfite sequencing and analysis

The same source of DNA used for PacBio HiFi long-read genome sequencing was used for whole-genome bisulfite sequencing. Whole-genome sequencing libraries were generated from 1ug of genomic DNA spiked with 0.1% (w/w) unmethylated λ DNA (Promega) previously fragmented to 300–400 bp peak sizes using the Covaris focused-ultrasonicator E210. Fragment size was controlled on a Bioanalyzer DNA 1000 Chip (Agilent) and the KAPA High Throughput Library Preparation Kit (KAPA Biosystems) was applied. End repair of the generated dsDNA with 3′- or 5′-overhangs, adenylation of 3′-ends, adaptor ligation and clean-up steps were carried out as per KAPA Biosystems’ recommendations. The cleaned-up ligation product was then analyzed on a Bioanalyzer High Sensitivity DNA Chip (Agilent) and quantified by PicoGreen (Life Technologies). Samples were then bisulfite converted using the EZ DNA Methylation-Lightning Gold Kit (Zymo), according to the manufacturer’s protocol. Bisulfite-converted DNA was quantified using OliGreen (Life Technologies) and, based on quantity, amplified by 9–12 cycles of PCR using the Kapa Hifi Uracil+DNA polymerase (KAPA Biosystems), according to the manufacturer’s protocol. The amplified libraries were purified using Ampure Beads and validated on Bioanalyzer High Sensitivity DNA Chips and quantified by PicoGreen. Libraries were sequenced on the Illumina NovaSeq6000 system using 150-bp paired-end sequencing. Whole genome bisulfite sequence reads were pre-processed with fastp to trim adapter and low-quality bases, then alignment was performed with Illumina’s DRAGEN aligner followed by post-processing with samtools, Picard, Bismark ^33^ and Bis-SNP ^34^ for marking of duplicates, methylation calling and SNV calling. To avoid potential biases in downstream analyses, we applied our benchmark filtering criteria as follows; ≥ 5 total reads, no overlap with SNPs (dbSNP 137), ≤ 20% methylation difference between strands, no overlap with DAC Blacklisted Regions (DBRs) or Duke Excluded Regions (DERs) generated by the ENCODE project: (http://hgwdev.cse.ucsc.edu/cgi-bin/hgFileUi?db=hg19&g=wgEncodeMapability). Methylation values at each site were calculated as total (forward and reverse) non-converted C-reads over total (forward and reverse) reads. CpGs were counted once per location combining both strands together.

### Correlation of CpG methylation across platforms

For sample-wise correlation (N=93), CpGs sequenced at >20x coverage across both platforms were selected and used to measure the linear correlation using Pearson correlation coefficient (r). For CpG-based correlations, the HiFi-GS data set was used to select the 500 CpGs with the highest standard deviation (SD) for the methylation rate. From the set of 93 samples with both HiFi-GS and WGBS, the methylation rates at these 500 CpGs were extracted. Then, for each CpG, the correlation between the HiFi-GS methylation rate and the WGBS methylation rate was computed using Pearson correlation coefficient (r). Also, for each CpG, a control correlation rate was computed by shuffling the methylation rates of all 93 samples and then computing the correlation between the HiFi-GS methylation rate and the (now-shuffled) WGBS methylation rate, whenever a matching methylation rate was available.

### Classification of rare extreme hypermethylation outliers

The HiFi-GS data set was used to calculate mean, standard deviation (SD) and 95th quantile (Q95) for each of the measured CpGs (**Supplementary Table 2)**. Then, for each HiFi-GS sample, the methylation rate and z-score for each CpG at 10x coverage was computed using all HiFi reads sequenced. This was repeated for each sample for each CpG at 10x read coverage using only reads phased to haplotype 1 (hap1) and repeated a third time using only the reads phased to haplotype 2 (hap2). Consequently, methylation rate and z-score were obtained for each combination of individual, CpG chromosomal position and phasing mode (all, hap1 or hap2) whenever the corresponding read depth was 10 or greater. A CpG position for an individual and phasing mode (all, hap1 or hap2) was flagged as hypermethylated if the methylation rate deviated three SD higher than the Q95 for that CpG. A CpG position for and individual and phasing mode was flagged as hypomethylated if deviating three SD lower than the Q5 for that CpG. Then, all CpGs for an individual and phasing mode were grouped into 200bp tiles across all autosomes. A 200bp tile for an individual and phasing mode was identified as an extreme hyper-mCpG tile if it contained two or more hypermethylated CpGs and no hypomethylated CpGs. Extreme hyper-mCpG tiles were further reported as rare in two ways: 1) hyper-mCpG tiles at the same position from any phasing mode are present in two or fewer unrelated individuals, reporting the hyper-mCpG tiles for an individual when the average methylation rate z-score is greater than 2 across all CpGs for the hyper-mCpG tile of that individual for a phasing mode; or 2) hyper-mCpG tiles at a position are potentially present in three or more unrelated individuals, but only hyper-mCpG tiles with an average methylation rate z-score greater than 5 across the CpGs in the hyper-mCpG tile of that individual for a phasing mode was kept. If the hyper-mCpG tile was present across multiple unrelated individuals, a minimum separation of three z-score units across the unrelated samples was required allowing only a maximum of two samples per tile.

The hyper-mCpG tiles were annotated with the closest OMIM gene position, the closest gene transcription start size (TSS) and overlapping DNase I hypersensitivity sites (DHS)^35^. For enrichment analysis of extreme hyper-mCpG tiles in regulatory element annotated by DHS, two sets of randomly selected hyper-mCpG tiles were created requiring the tile being present in at least 5 samples (out of 276) and with a mean methylation rate z score per tile corresponding -1 <= z <= 1 and -2 <= z <= 2, respectively. These criteria resulted in a total of 10,129,897 and 10,134,018 hyper-mCpG tiles, respectively, of which 50,000 were randomly selected from each set and used as background control.

### Mapping genetic variation to extreme hypermethylation outliers

The distance from the extreme bin to the closest gnomAD ^36^ high confidence variant in the sample with the extreme bin was compared to the distance from the same bin coordinates to the closest gnomAD high confidence variant in all other samples (with a variant on that chromosome) to obtain a percentile rank. The distance to the closest phased (via WhatsHap) heterozygote gnomAD high confidence variant on hap1 in the sample was also compared with the extreme bin against the distance from that bin coordinate to the closest phased hap1 gnomAD variant in all other samples to obtain a percentile rank, and similarly for the closest phased heterozygote variant on hap2.

The excess familial sharing of hypermethylation outliers was observed to be significant in random permutations among samples that had family members available. Specifically, among called hypermethylation outliers (max two called per window) there were total 2,465 observations in samples with related individual among to cohort, of these 290 were shared with the related family member (11.8%). The familial relationships (family labels) were then permuted in the hypermethylation tiles and calculated random overlap of familial sharing observing on average 94.2 (range 36-58 per permutation run) such overlaps by chance indicating an 3.079x excess of familial sharing (290/94.2).

### PacBio HiFi long-read transcript (isoform) sequencing and analysis

RNA was isolated from 17 iPSC samples using a RNeasy Mini Kit (Qiagen, Cat. No. 74104), following the Quick Start Protocol associated with the kit. The RNA concentration for each sample was determined with a Qubit RNA BR Assay Kit (ThermoFisher, Q10210) and the RIN was determined with a RNA ScreenTape (Agilent, Cat. No. 5067-5577 and 5067-5576) on the TapeStation platform. A maximum of 300ng of RNA with a RIN score greater than 7.0 was aliquoted from each sample to be used as an input into cDNA synthesis for Iso-Seq library preparation. If the concentration of RNA was too low for there to be 300ng of RNA in 7μL, 300ng of RNA was aliquoted into a 1.5mL tube and then the sample was concentrated using a vacuum concentration system without heating until the sample was less than or equal to 7μL in volume. The RNA underwent cDNA synthesis with a NEBNext Single Cell/Low Input cDNA Synthesis & Amplification Module (NEB, Cat. No. E6421S) and an Iso-Seq Express Oligo Kit (PacBio, Cat. No. 101-737-500) as described in the Iso-Seq Express Template Preparation for Sequel and Sequel II Systems protocol from PacBio. The samples were not amplified with barcoded primers during the cDNA amplification steps since the downstream libraries would not be multiplexed for sequencing. The amplified cDNA was purified for long transcripts greater than 3kb in length after the first cDNA amplification step. The quantity of cDNA was determined with a Qubit dsDNA HS Assay Kit (ThermoFisher, Cat. No. Q32854) and if the cDNA yield was less than 160ng, the required input for the Sequel II system, the samples were reamplified following the procedure described in Appendix 1 of the Iso-Seq protocol. NEBNext High-Fidelity 2X PCR Master Mix (NEB, Cat. No. M0541S) used in the PCR reamplification master mix and after reamplification, the cDNA was bead-cleaned following the low-yielded sample procedure. The cDNA was quantified again to ensure that there was adequate yield for SMRTbell library preparation. In total, 160 to 500ng of cDNA from each sample was used as an input into the SMRTbell library preparation protocol without pooling the cDNA. The final concentration of the libraries was determined with a Qubit dsDNA HS Assay Kit and the library size was determined with a High Sensitivity D5000 ScreenTape (Agilent, Cat. No. 5067-5592 and 5067-5593) on the TapeStation platform. Libraries were sequenced on the Sequel IIe Systems using the Sequel II Binding Kit 2.0 (101-842-900) or 2.1 (101-820-500) and Sequel II Sequencing Kit 2.0 (101-820-200) with 30 hr movies/SMRT cell at 90pM loading with 1 cell/sample. The IsoSeq3 pipeline (https://github.com/PacificBiosciences/IsoSeq) was used to generate full-length non-concatemer (FLNC) reads which were then aligned to the reference genome (GRCh38) using minimap2 with the argument -ax splice:hq. Reads overlapping a CCDS region and informative of a heterozygous SNV were kept and each allele was count for each read covered by at least 5X.

### Clinical Validation

“Short” PCR amplification of the normal allele of the FRA12A/*DIP2B* repeat– containing region was performed with the aid of 2X Failsafe buffer J (received with FailSafe enzyme) with use of primers derived from the sequences flanking the repeat (DIP2B-CGG-F primer, 5’ - GTCTTC[1]AGCCTGACTGGGCTGG-3’, and DIP2B-CGG-R, primer 5’-CCGG[1]CGACGGCTCCAGGCCTCG-3’. 95°C-3’; (95°C-1’;60°C-1’-0.5/cycle; 72°C-1’)x10; (95°C-1’; 55°C-1’;72-1’)x25; 72°C-1’, 4°C ∞). The PCR products were electrophoresed on a 1.5% agarose gel. Triplet-repeat PCR was also performed by primed PCR according to the principles described previously ^37,38^ to detect CAG-repeat expansion.

Three different primers were added to the PCR mixture: a single forward fluorescently labeled primer (DIP2B-CGG-F) and a combination of two reverse primers (P4, 5’ - TACGCATCCCAGTTTGAGACGGCCGCCGCCGCCGCCGC-3’, and P3, 5’ - TACGCATCCCAGTTTGAGACG-3’) in a 1:10 ratio. The reverse primer P4 anneals at different sites of the CGG repeat, which produces PCR products of different lengths that differ from each other by a multiple of three residues. After depletion of the P4 primer, the P3 primer takes over and amplifies the PCR products of different lengths. TP-PCR conditions (95-7’ (95-1”, 63+0.5/cycle-1”, 72-1’) x10, (95-30”, 56-30”, 72-1’+10”/cycle) x25, 72-10’,4C-∞) Diluted PCR products (1:5) were mixed with 1200LIZ and Hi-Di Formamide, PCR products were size fractionated on a Prism ABI 3500 DNA sequencer (Applied Biosystems).

## Data Availability

All data produced in the present study are available upon request via dbGAP using accession number phs002206.v4.p1

## Data availability

All data are deposited to dbGAP (https://www.ncbi.nlm.nih.gov/gap/) under the accession number phs002206.v4.p1.

## Code availability

Only publicly available tools were used in data analysis as described wherever relevant in Methods.

## Acknowledgment

We would like to thank all families for participating in our study. This work was made possible by the generous gifts to Children’s Mercy Research Institute and Genomic Answers for Kids program at Children’s Mercy Kansas City. We also would like to thank Nick Nolte, Dan Louiselle and Rebecca Biswell for their work in sample processing; Laura Puckett and Adam Walters for their work in library preparation and sequencing and the GA4K coordination team led by Bradley Belden for their work in clinical coordination. T.P holds the Dee Lyons/Missouri Endowed Chair in Pediatric Genomic Medicine and E.G holds the Roberta D. Harding & William F. Bradley, Jr. Endowed Chair in Genomic Research.

## Ethical declarations

The study was approved by the Children’s Mercy Institutional Review Board (IRB) (Study # 11120514). Informed written consent was obtained from all participants prior to study inclusion.

