## Supplementary Figures for "Direct haplotype-resolved 5-base HiFi sequencing for genome-wide profiling of hypermethylation outliers in a rare disease cohort"

**This PDF file includes:**
Supplementary Figures 1-20

**Other supplementary materials for this manuscript include the following:**
Supplementary Tables (separate excel file)

**Supplementary Figure 10…………………………………………………………...Page 12**
**Supplementary Figure 11…………………………………………………………...Page 13**
**Supplementary Figure 12…………………………………………………………...Page 14**
**Supplementary Figure 13…………………………………………………………...Page 15**
**Supplementary Figure 14…………………………………………………………...Page 16**
**Supplementary Figure 15…………………………………………………………...Page 17**
**Supplementary Figure 16…………………………………………………………...Page 18**
**Supplementary Figure 17…………………………………………………………...Page 19**
**Supplementary Figure 18…………………………………………………………...Page 20**
**Supplementary Figure 19…………………………………………………………...Page 21**
**Supplementary Figure 20…………………………………………………………...Page 22**

**Supplementary Figure 1.** Density plot showing the distribution of CpG-based correlation estimates (Pearson R, x-axis) of the top (N=500) variable CpGs obtained from 93 samples profiled by HiFi-GS and WGBS. Blue line represents CpG-based measures of the 500 tested CpGs and red line represents similar number but when WGBS values are permuted.

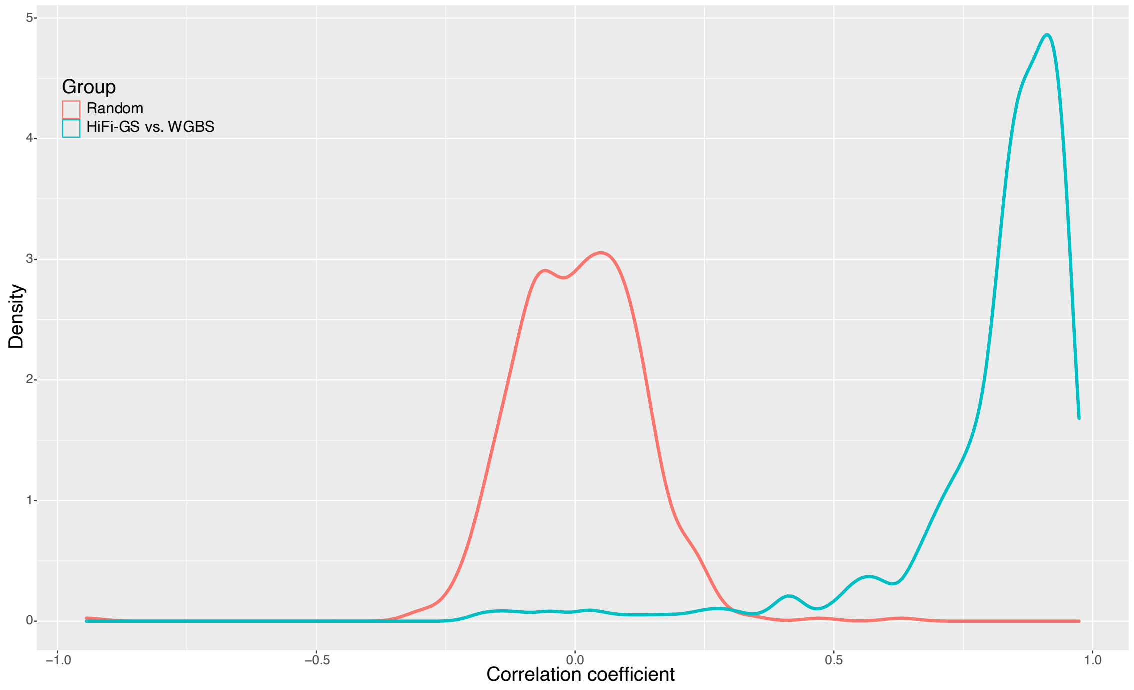

**Supplementary Figure 2.** Haplotype-resolved HiFi-GS of an imprinted region (chr20: 58,851,237-58,851,799) at the *GNAS* locus associated with Albright hereditary osteodystrophy for a complete trio showing maternal allele-specific hypermethylation in proband (black boxes). Haplotype-resolved HiFi-GS reads are depicted with CpG modification staining (blue indicating low CpG methylation prediction and red indicating high CpG methylation prediction). Hap 1 denotes haplotype 1 and Hap 2 denotes haplotype 2.

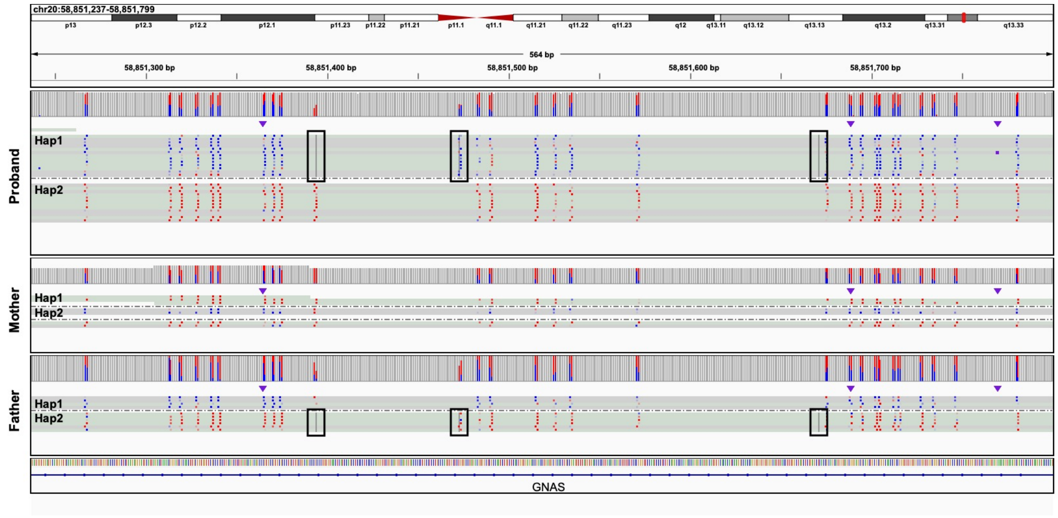

**Supplementary Figure 3.** Haplotype-resolved HiFi-GS of an imprinted region (chr6:144,008,510-144,008,709) at the *PLAGL1* locus associated with transient neonatal diabetes mellitus showing maternal allele-specific hypermethylation in proband (black boxes). Haplotype-resolved HiFi-GS reads are depicted with CpG modification staining (blue indicating low CpG methylation prediction and red indicating high CpG methylation prediction). Hap 1 denotes haplotype 1 and Hap 2 denotes haplotype 2.

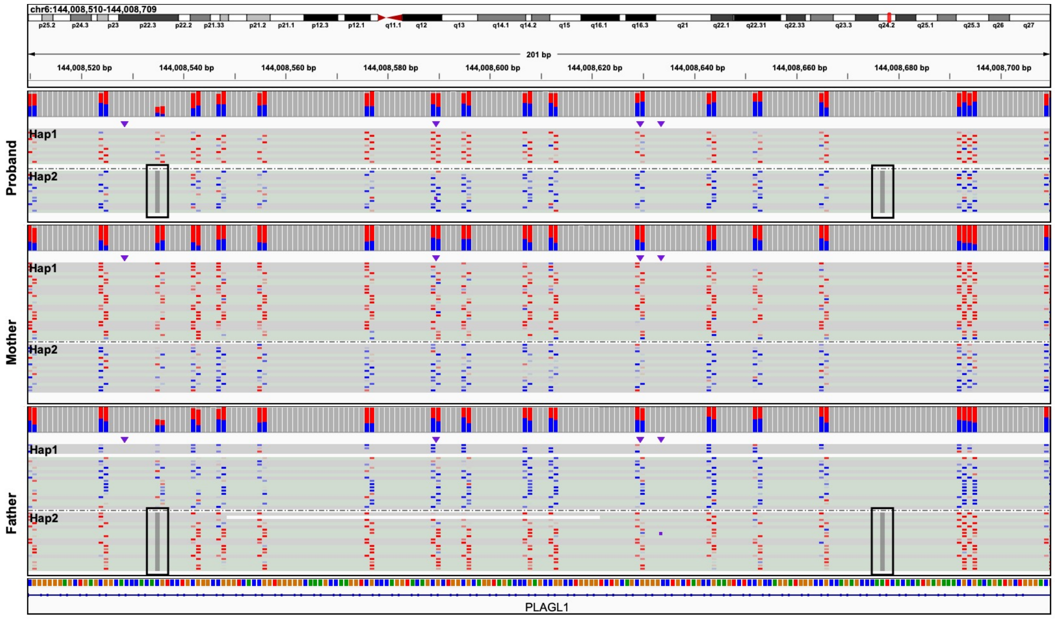

**Supplementary Figure 4.** Haplotype-resolved HiFi-GS of an imprinted region (chr15: 23,648,164-23,648,386) at the *MAGEL2* locus associated with Schaaf-Yang syndrome showing maternal allele-specific hypermethylation in proband (black boxes). Haplotype-resolved HiFi-GS reads are depicted with CpG modification staining (blue indicating low CpG methylation prediction and red indicating high CpG methylation prediction). Hap 1 denotes haplotype 1 and Hap 2 denotes haplotype 2.

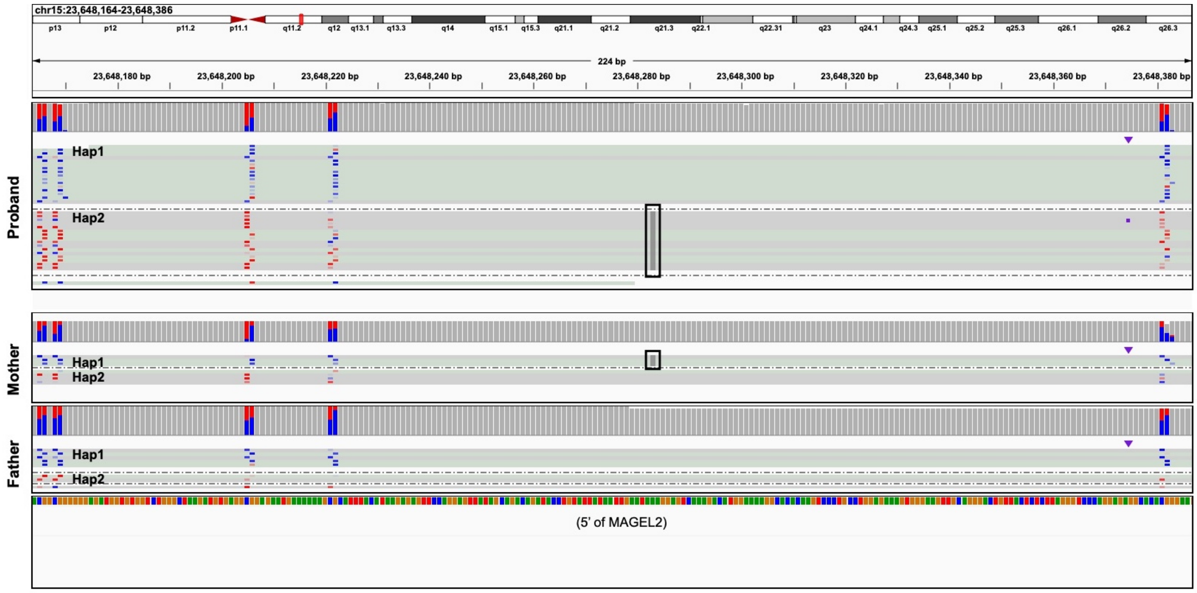

**Supplementary Figure 5.** Haplotype-resolved HiFi-GS of an imprinted region (chr14: 100,809,369-100,809,835) at the *MEG3*/*DLM1* locus associated with Temple syndrome showing paternal allele-specific hypermethylation in proband (black boxes). Haplotype-resolved HiFi-GS reads are depicted with CpG modification staining (blue indicating low CpG methylation prediction and red indicating high CpG methylation prediction). Hap 1 denotes haplotype 1 and Hap 2 denotes haplotype 2.

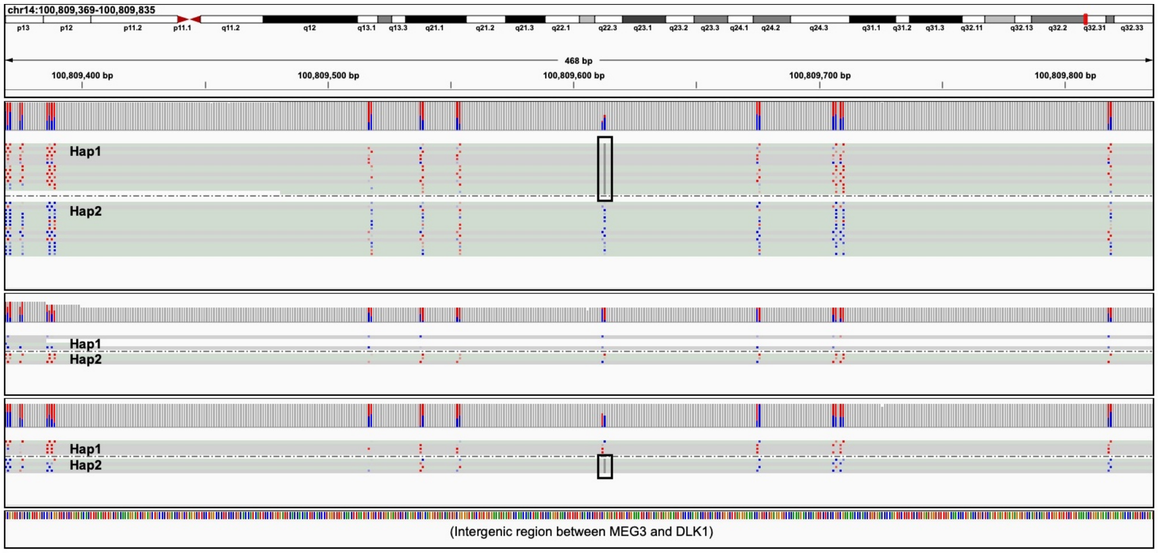

**Supplementary Figure 6.** Validation of HiFi-GS resolved allele-specific methylation by WGBS. Genomic view of an example of a rare SNV (black box) mapping in *cis* close to a hyper-mCpG tile on chromosome 10 causing allele-specific hypermethylation. Track depicts haplotype-resolved HiFi-GS reads with CpG modification staining (blue indicating low methylation prediction and red indicating high methylation prediction). Hap 1 denotes haplotype 1 and Hap 2 denotes haplotype 2. Top tracks depict parallel assessment of CpG methylation (y-axis, 0-100%) by WGBS in the proband and an unrelated control sample not carrying the rare SNV.

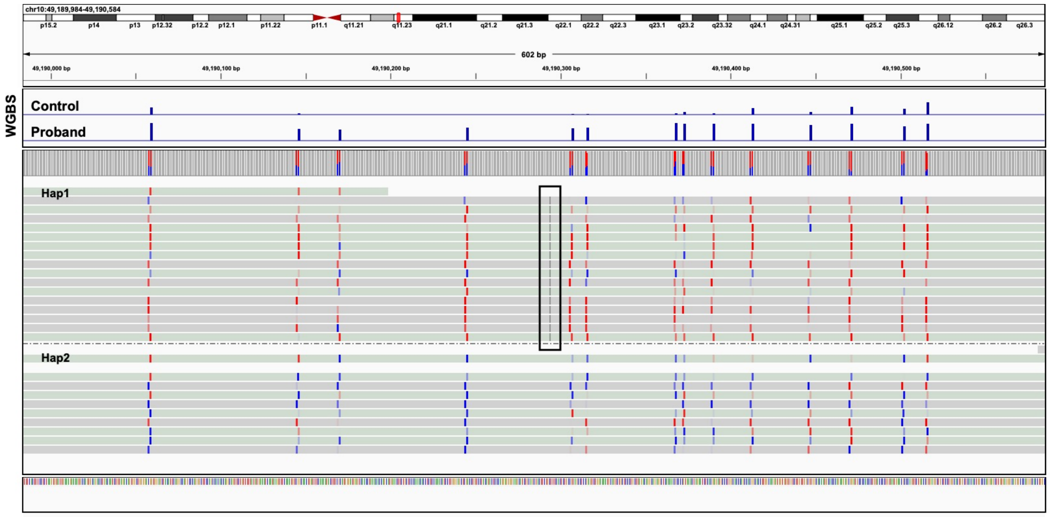

**Supplementary Figure 7.** Validation of HiFi-GS resolved allele-specific methylation by WGBS. Genomic view of an example of a rare SNV (black box) mapping in *cis* close to a hyper-mCpG tile on chromosome 22 causing allele-specific hypermethylation. Track depicts haplotype-resolved HiFi-GS reads with CpG modification staining (blue indicating low methylation prediction and red indicating high methylation prediction). Hap 1 denotes haplotype 1 and Hap 2 denotes haplotype 2. Top tracks depict parallel assessment of CpG methylation (y-axis, 0-100%) by WGBS in the proband and an unrelated control sample not carrying the rare SNV.

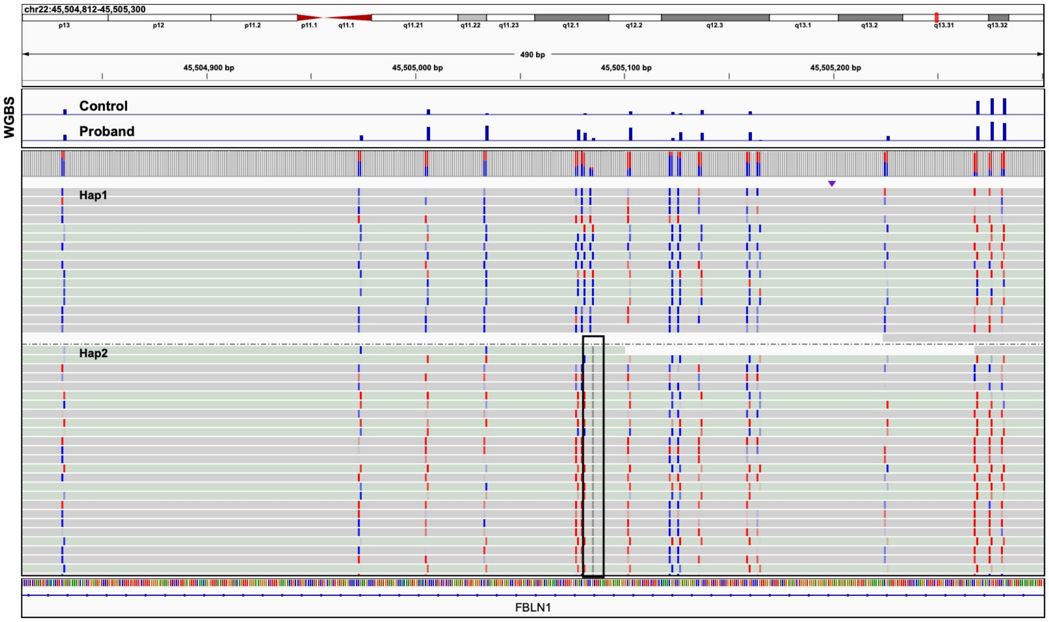

**Supplementary Figure 8.** Validation of HiFi-GS resolved allele-specific methylation by WGBS. Genomic view of an example of a rare SNV (black box) mapping in *cis* close to a hyper-mCpG tile on chromosome 10 causing allele-specific hypermethylation. Track depicts haplotype-resolved HiFi-GS reads with CpG modification staining (blue indicating low methylation prediction and red indicating high methylation prediction). Hap 1 denotes haplotype 1 and Hap 2 denotes haplotype 2. Top tracks depict parallel assessment of CpG methylation (y-axis, 0-100%) by WGBS in the proband and an unrelated control sample not carrying the rare SNV.

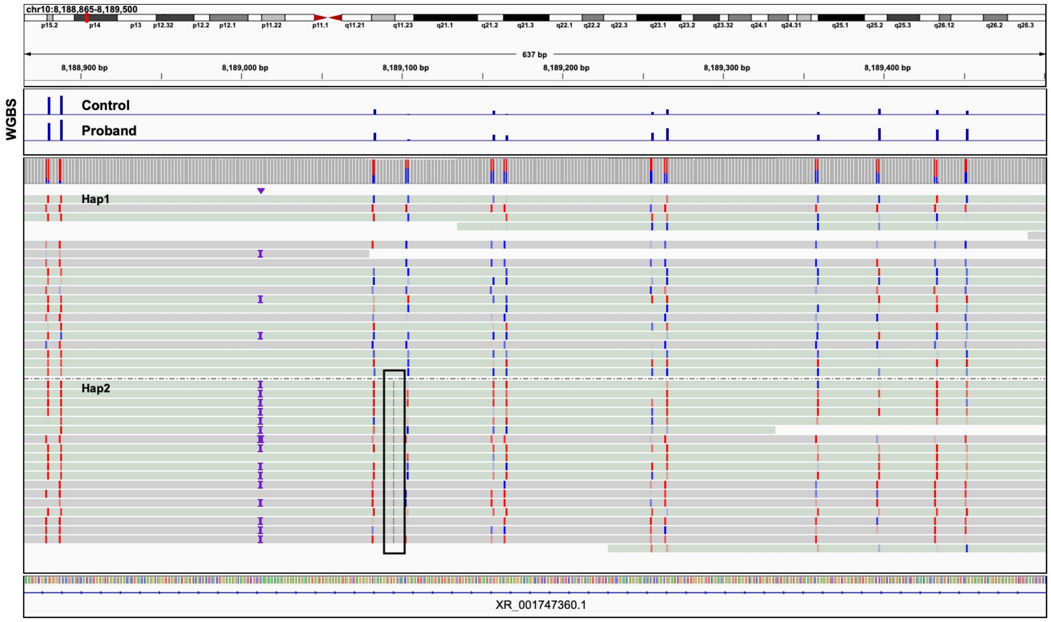

**Supplementary Figure 9**. Genomics view of 160 bp in an intergenic region comprising an 8 bp deletion (Proband 1 Hap1) resolved by HiFi-GS that results in proximal hypermethylation. Track depicts haplotype-resolved HiFi-GS reads with CpG modification staining (blue indicating low methylation prediction and red indicating high methylation prediction). Hap 1 denotes haplotype 1 and Hap 2 denotes haplotype 2.  Top tracks depict parallel assessment of CpG methylation (y-axis, 0-100%) by WGBS in the proband and an unrelated control sample not carrying the deletion.

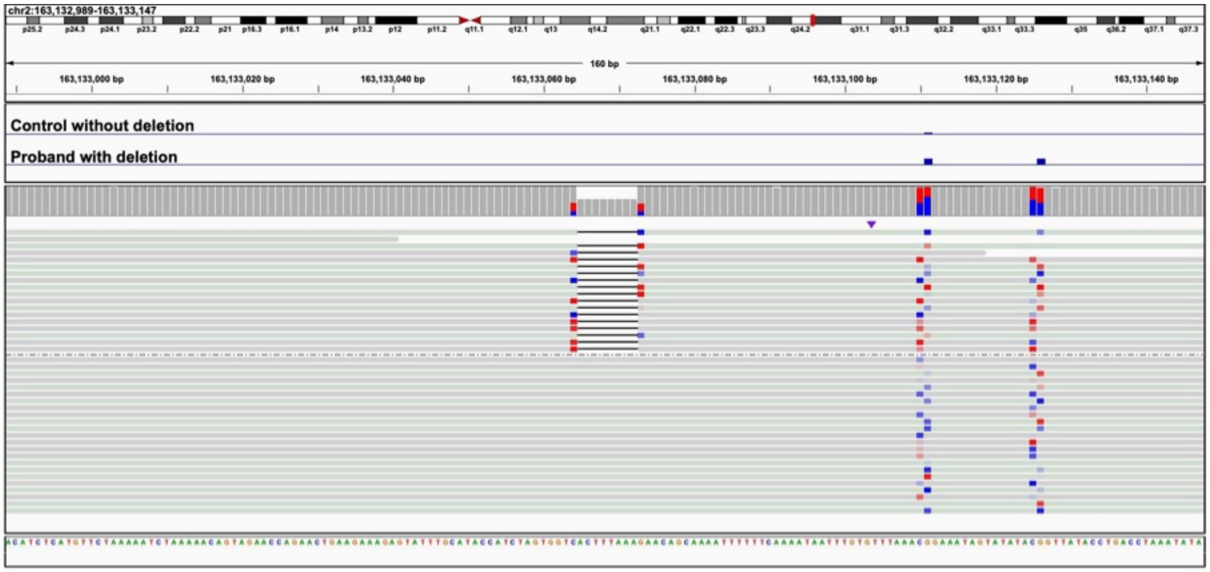

**Supplementary Figure 10.** For the autosomal recessive disease gene *NHLRC2*, an A to C transversion (chr10:113,854,859) maps 900bp upstream of a rare hyper-mCpG tile (red box). To assess the size (in base pair) of the extreme hyper-CpG tile linked to the rare, local (*cis*) variant, HiFi-GS reads (bottom tracks) overlapping the candidate *cis*-variant were fetched. Track depicts haplotype-resolved HiFi-GS reads with CpG modification staining (blue indicating low methylation prediction and red indicating high methylation prediction). Top track depicts P-values (-log10, 0-11) from 2-by-2 Fisher's exact test examining each CpG state (methylated vs. unmethylated) in reads with rare C versus common A-allele carrying reads, respectively. Hypermethylation shows statistical significance for the phased C-allele from haplotype-resolved HiFi-GS data across ~200-300bp. Second and third track from the top depict average CpG methylation level (y-axis, 0-100%) across all C carrying reads (rare) measured by HiFi-GS.

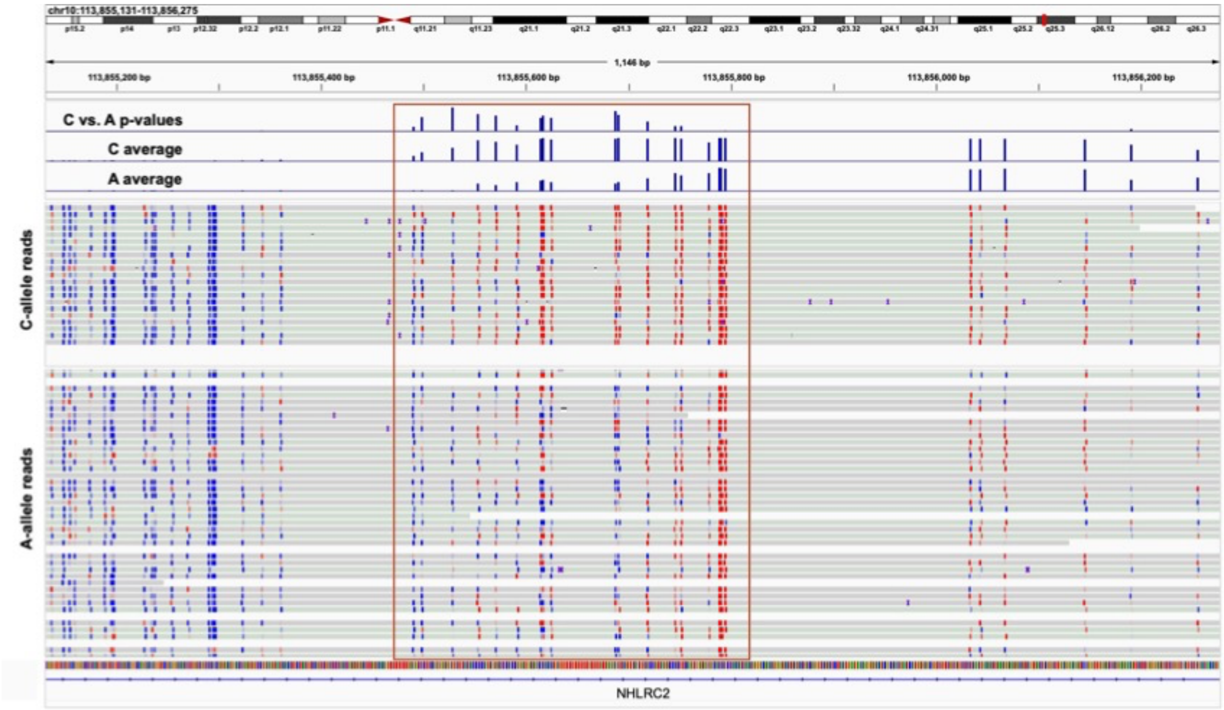

**Supplementary Figure 11. A**. Genomics view of 82 bp at an intergenic locus showing a rare SNV (blue box) that results in proximal hypermethylation in one allele (Hap2). Track depicts haplotype-resolved HiFi-GS reads with CpG modification staining (blue indicating low methylation prediction and red indicating high methylation prediction). Hap 1 denotes haplotype 1 and Hap 2 denotes haplotype 2. **B**. Zoomed out region in UCSC genome browser showing overlap of SNV (blue line) with *cis*-regulatory element (CRE) as mapped by ENCODE.

**A**

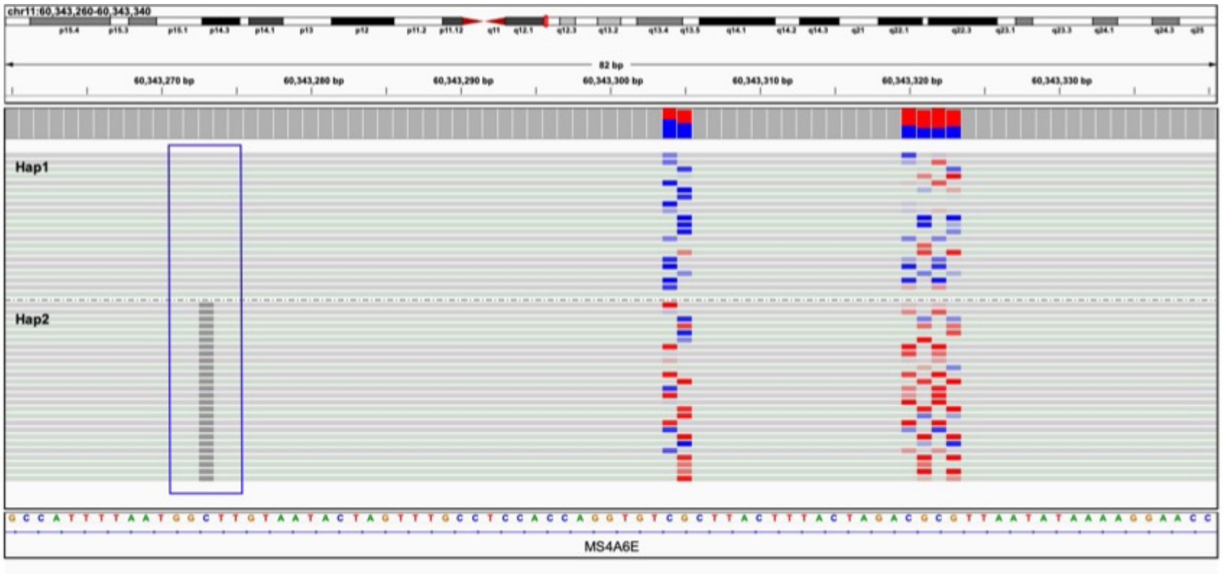

**B**

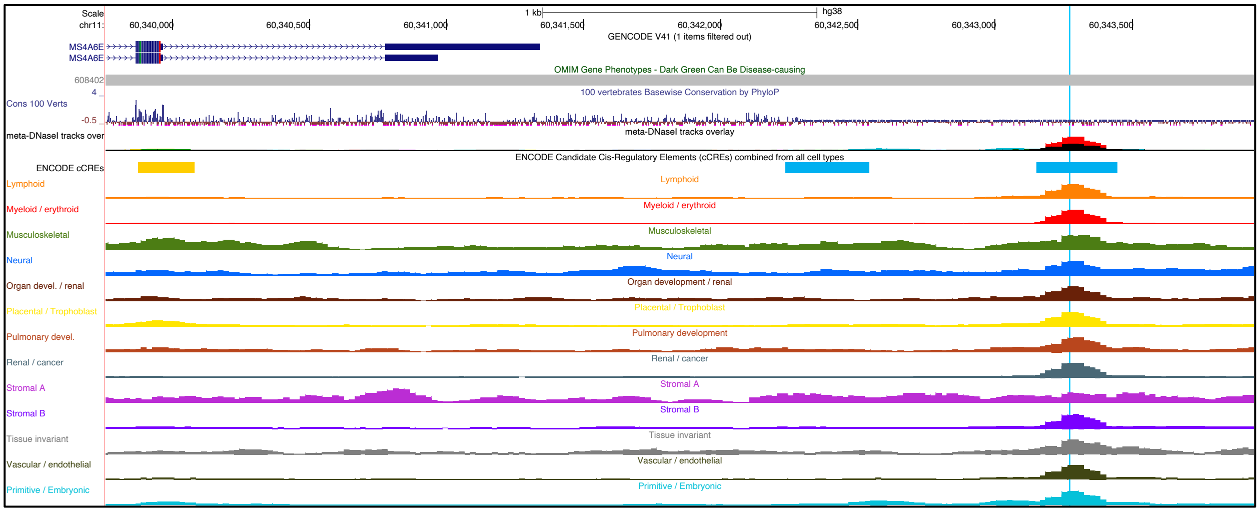

**Supplementary Figure 12. A.** Genomics view of 13kb at the *LOC199882* locus comprising a breakpoint of a CNV (black box, Proband 2) that results in proximal hypermethylation. Lower tracks depict haplotype-resolved HiFi-GS reads with CpG modification staining (blue indicating low methylation prediction and red indicating high methylation prediction) in a proband with and without the CNV. Hap 1 denotes haplotype 1 and Hap 2 denotes haplotype 2. Upper tracks depict CpG methylation by WGBS (0-100%, y axis) in the same samples confirming hypermethylation in proband carrying the CNV. **B.** Zoomed out region in UCSC genome browser showing overlap of CNV (blue box) with *cis*-regulatory element (CRE) as mapped by ENCODE.

**A**

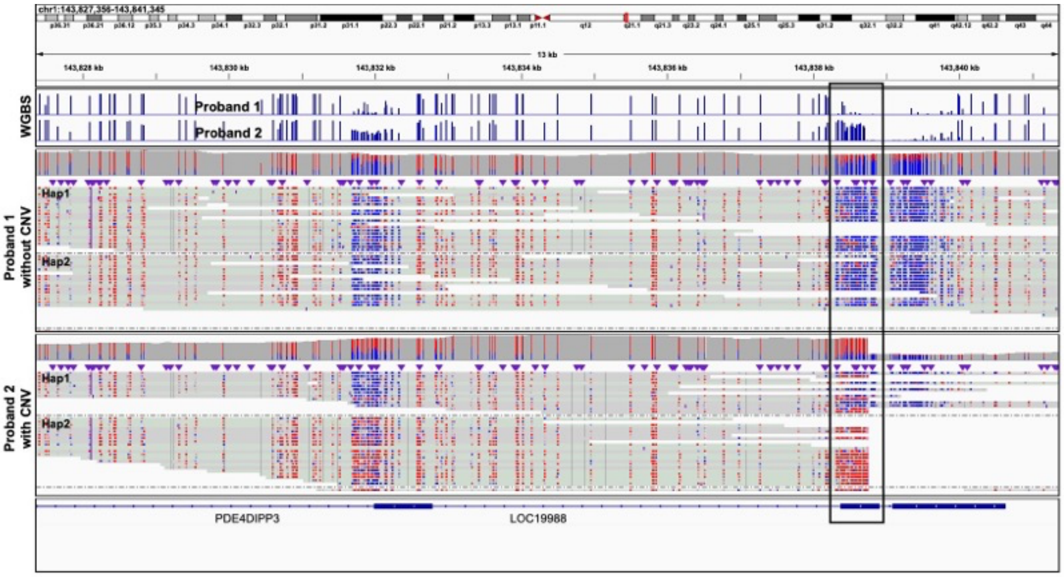

**B**

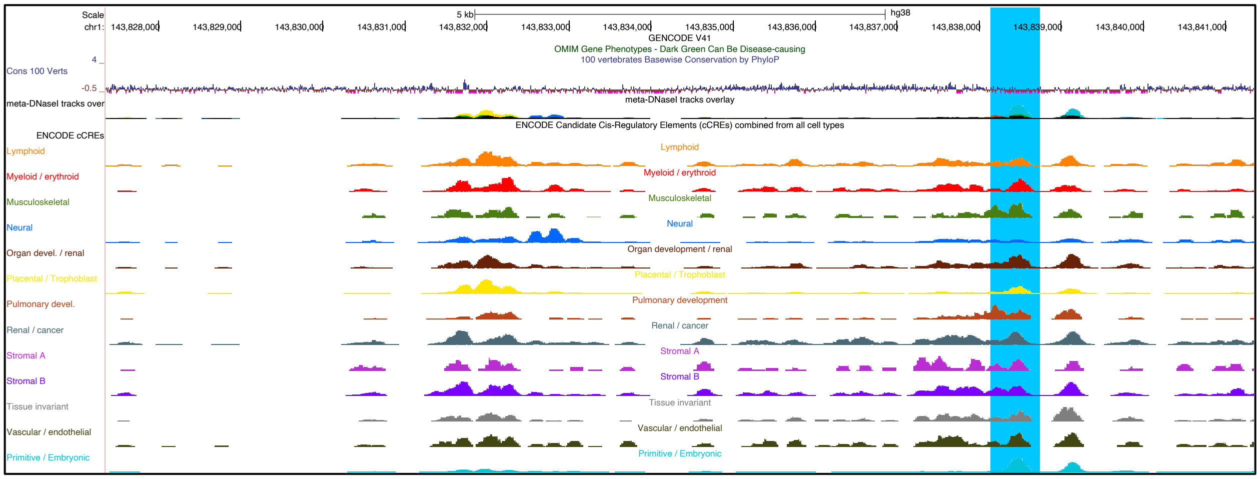

**Supplementary Figure 13. A.** Identification of a previously uncharacterized repeat expansion at *ELF1* locus causing hypermethylation. Tracks depict haplotype-resolved HiFi-GS reads with CpG modification staining (blue indicating low methylation prediction and red indicating high methylation prediction) in a proband with the repeat expansion. Hap 1 denotes haplotype 1 and Hap 2 denotes haplotype 2. **B.** Zoomed in region in UCSC genome browser showing overlap of repeat expansion (blue box) with *cis*-regulatory element (CRE) as mapped by ENCODE.

**A**

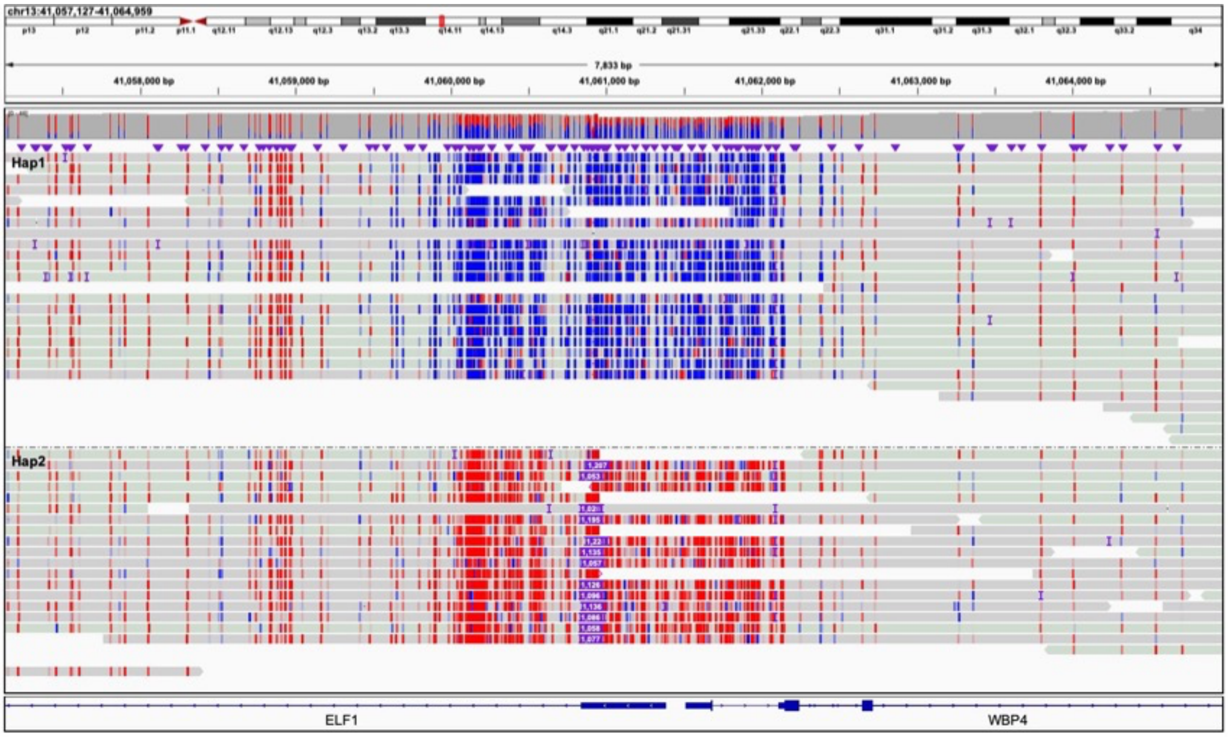

**B**

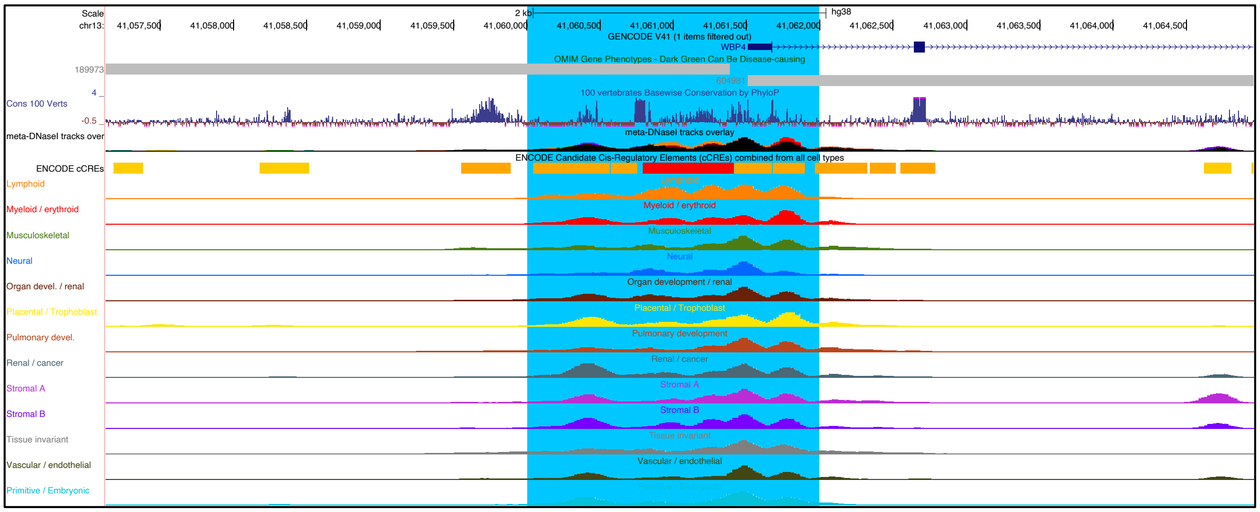

**Supplementary Figure 14. A.** Identification of a previously uncharacterized repeat expansion at *LINGO3* locus causing promoter hypermethylation. Tracks depict haplotype-resolved HiFi-GS reads with CpG modification staining (blue indicating low methylation prediction and red indicating high methylation prediction) in a proband with the repeat expansion. Hap 1 denotes haplotype 1 and Hap 2 denotes haplotype 2. **B.** Zoomed out region in UCSC genome browser showing overlap of repeat expansion (blue box) with *cis*-regulatory element (CRE) as mapped by ENCODE.

**A.**

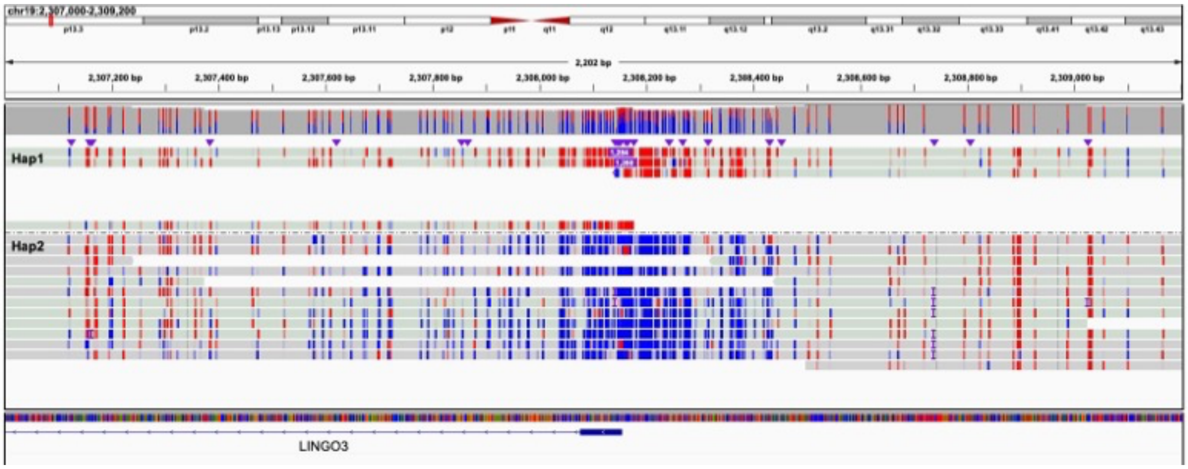

**B.**

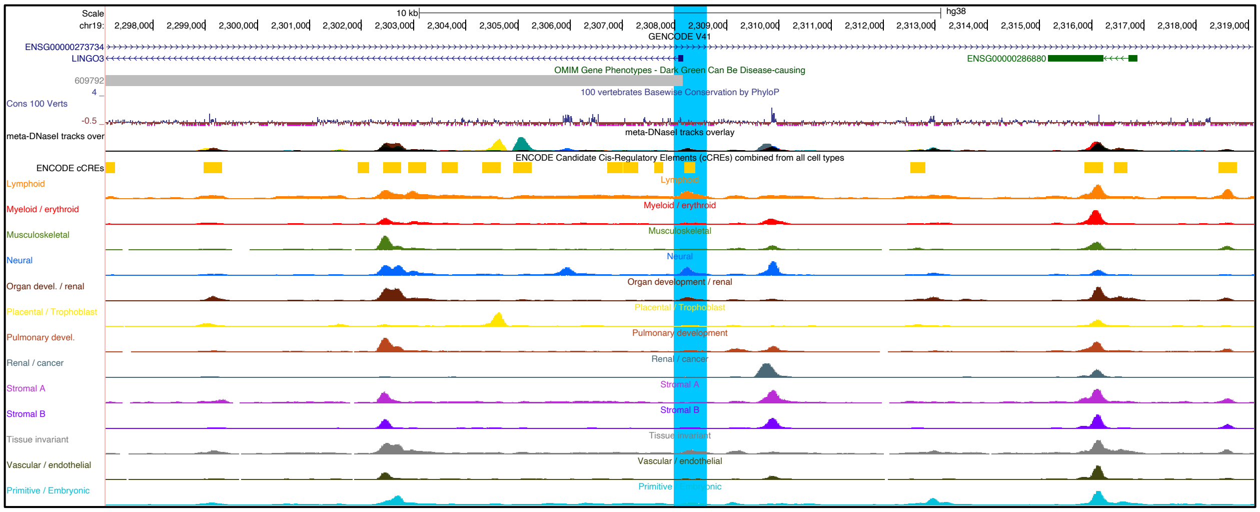

**Supplementary Figure 15. A.** Identification of a previously uncharacterized repeat expansion at *SNED1* locus causing hypermethylation. Tracks depict haplotype-resolved HiFi-GS reads with CpG modification staining (blue indicating low methylation prediction and red indicating high methylation prediction) in a proband with the repeat expansion. Hap 1 denotes haplotype 1 and Hap 2 denotes haplotype 2. **B.** Zoomed out region in UCSC genome browser showing overlap of repeat expansion (blue box) with *cis*-regulatory element (CRE) as mapped by ENCODE.

**A**

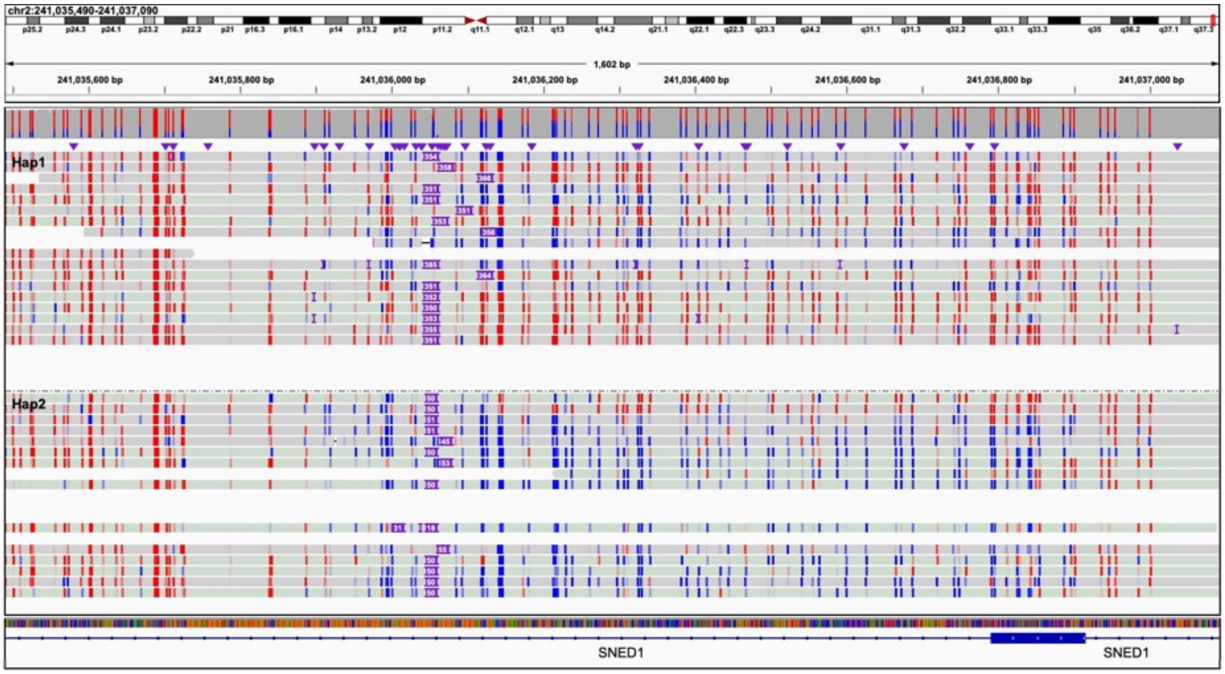

**B**

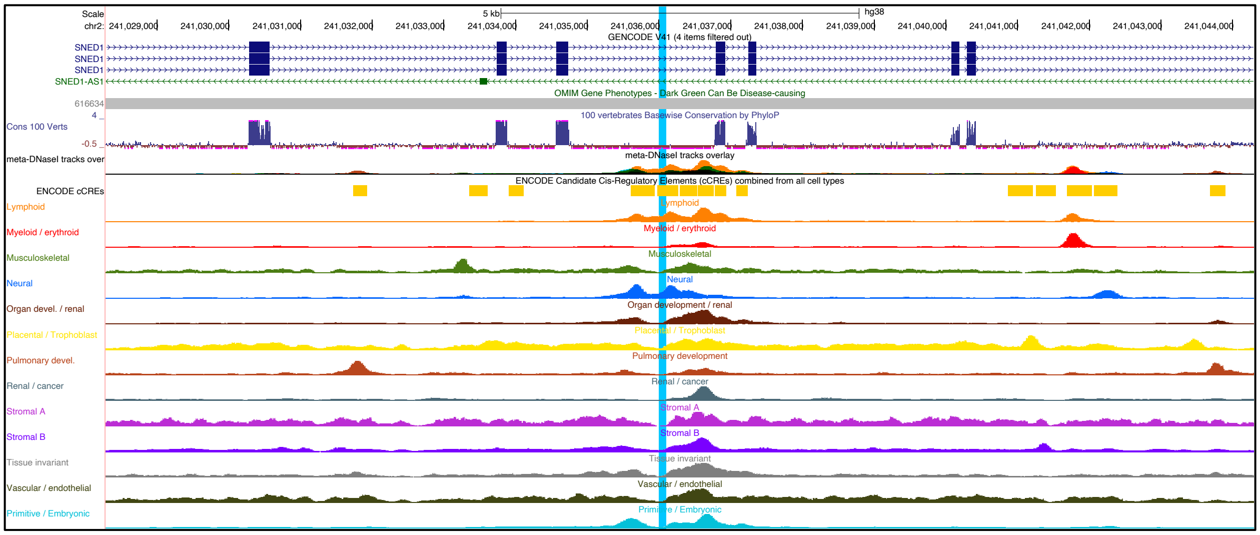

**Supplementary Figure 16. A**. Identification of a rare duplication (10.6kb, black box, solid line) causing "compensatory" promoter hypermethylation (black box, dashed line) at the *COX20* disease locus. Lower tracks depict haplotype-resolved HiFi-GS reads with CpG modification staining (blue indicating low methylation prediction and red indicating high methylation prediction) in complete trio showing maternal inheritance of the duplication in proband. Hap 1 denotes haplotype 1 and Hap 2 denotes haplotype 2. Top tracks show parallel assessment of CpG methylation by WGBS (y-axis, 0-100%) validating hypermethylation caused by the duplication. **B.** Zoomed in region in UCSC genome browser showing overlap of hypermethylation effect (grey box) caused by duplication (blue box) with *cis*-regulatory element (CRE) as mapped by ENCODE.

**A**

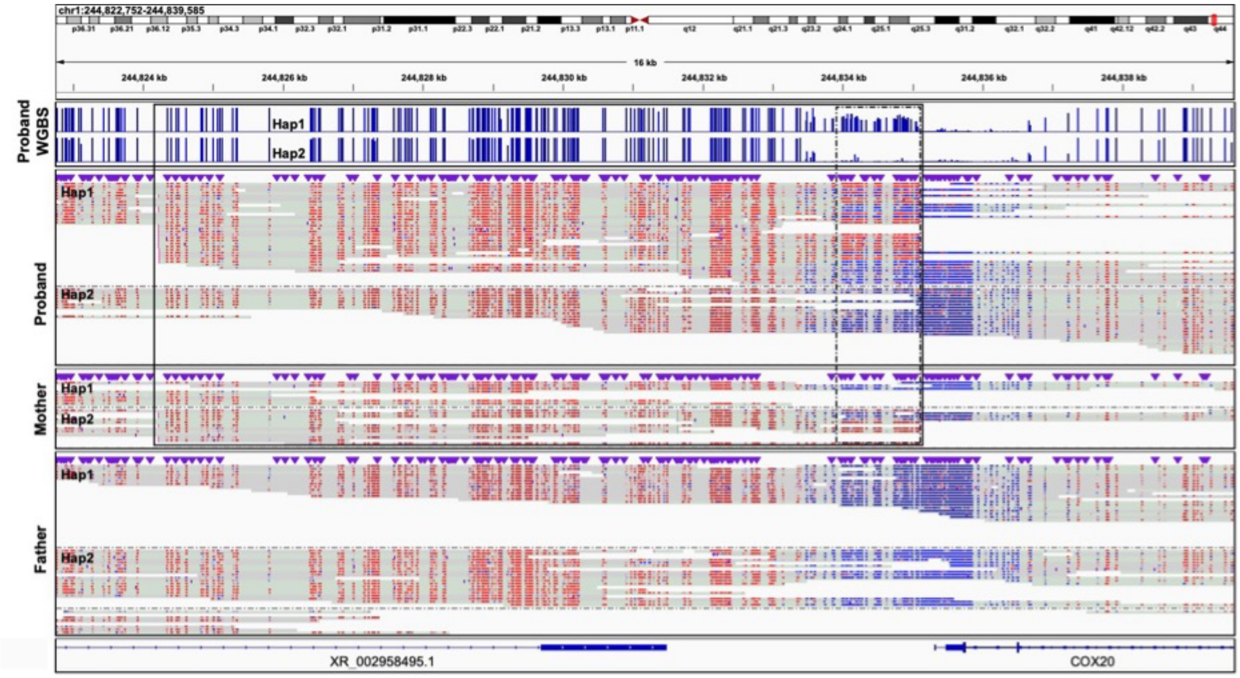

**B**

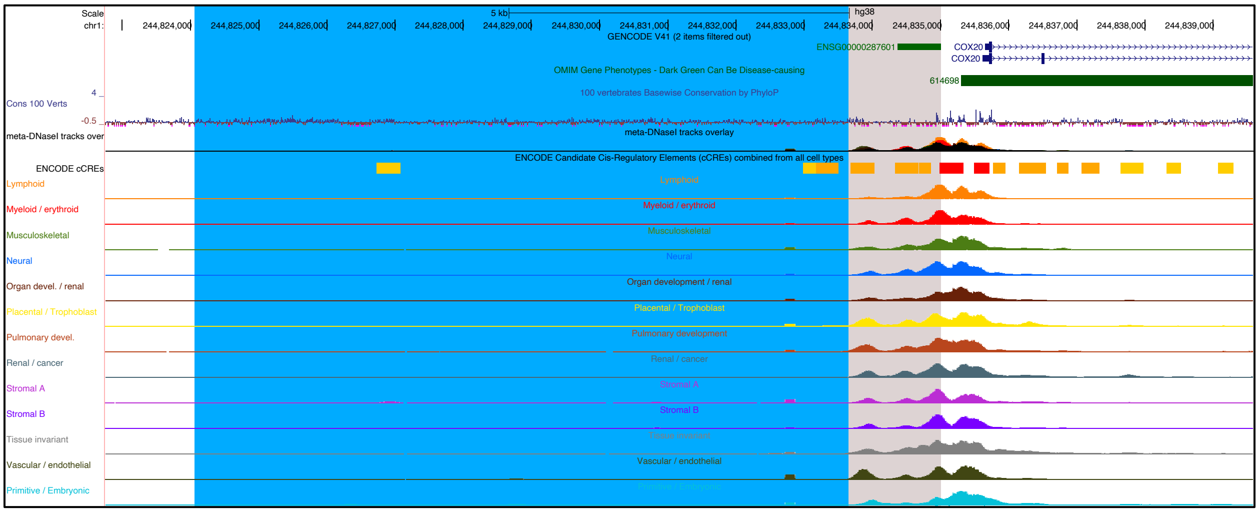

**Supplementary Figure 17. A.** Genomics view of ~3.5kb at the *RNF166* locus comprising an inherited deletion (Proband Hap2 and Mother Hap2) resolved by HiFi-GS that results in 2kb hypermethylation surrounding the deletion. Tracks depict HiFi-GS reads with CpG modification staining (blue indicating low methylation prediction and red indicating high methylation prediction) in proband and mother. Hap 1 denotes haplotype 1 and Hap 2 denotes haplotype 2. **B.** Zoomed in region in UCSC genome browser showing overlap of hypermethylation effect (light green box) caused by the deletion with *cis*-regulatory element (CRE) as mapped by ENCODE.

**A**

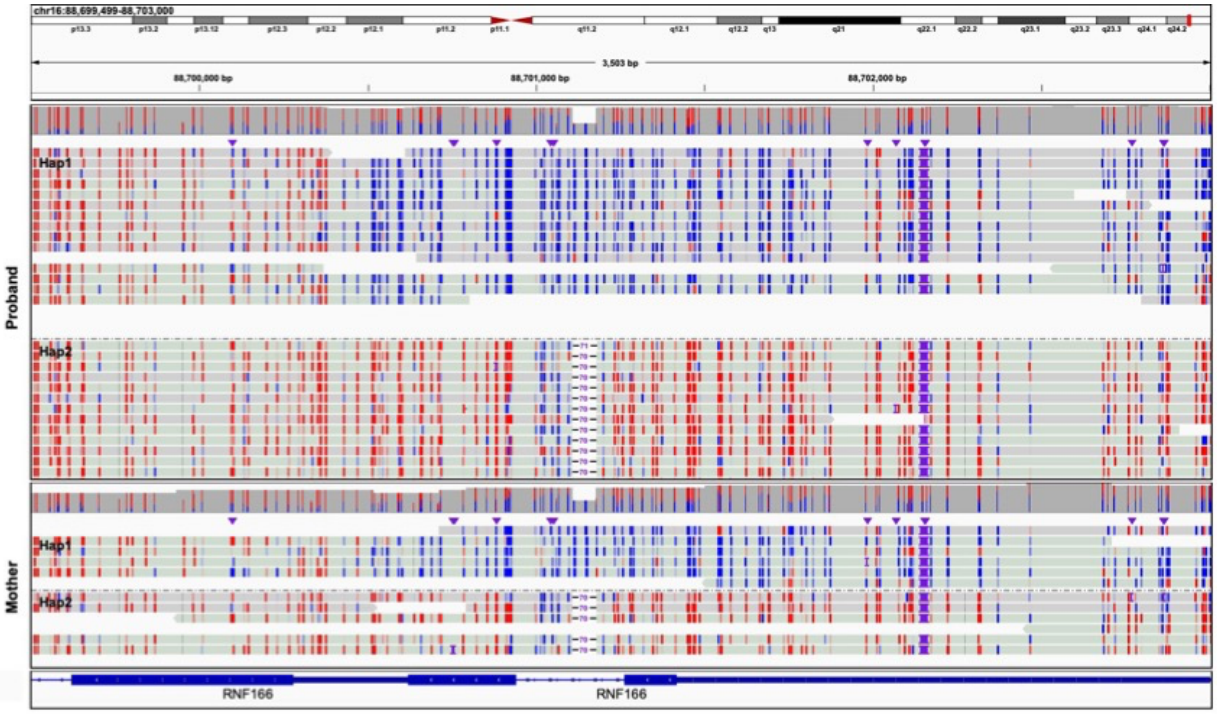

**B**

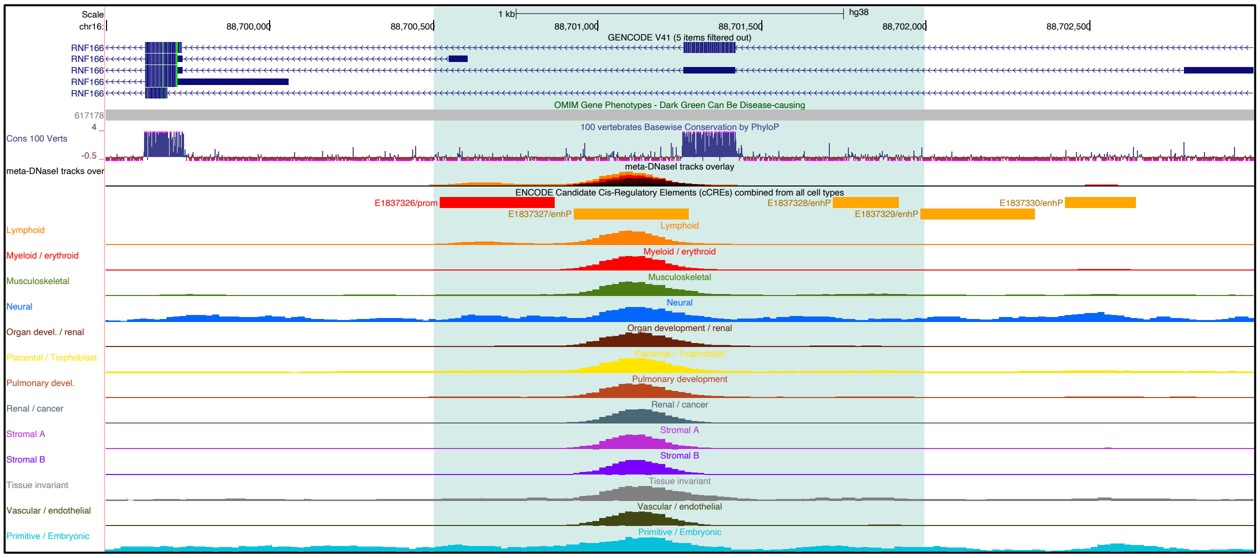

**Supplementary Figure 18. A.** Genomics view of ~300bp comprising a 13 bp deletion (blue box) shared in sibling (Sibling 1; Hap1 and Sibling 2; Hap2) that results in proximal hypermethylation. Lower tracks depict haplotype-resolved HiFi-GS reads with CpG modification staining (blue indicating low methylation prediction and red indicating high methylation prediction) in siblings and unrelated proband without the deletion. Hap 1 denotes haplotype 1 and Hap 2 denotes haplotype 2. Top tracks show parallel assessment of CpG methylation by WGBS (y-axis, 0-100%) validating hypermethylation caused by the deletion. **B.** Zoomed out region in UCSC genome browser showing overlap of hypermethylation effect (light green box) caused by deletion with *cis*-regulatory element (CRE) as mapped by ENCODE.

**A**

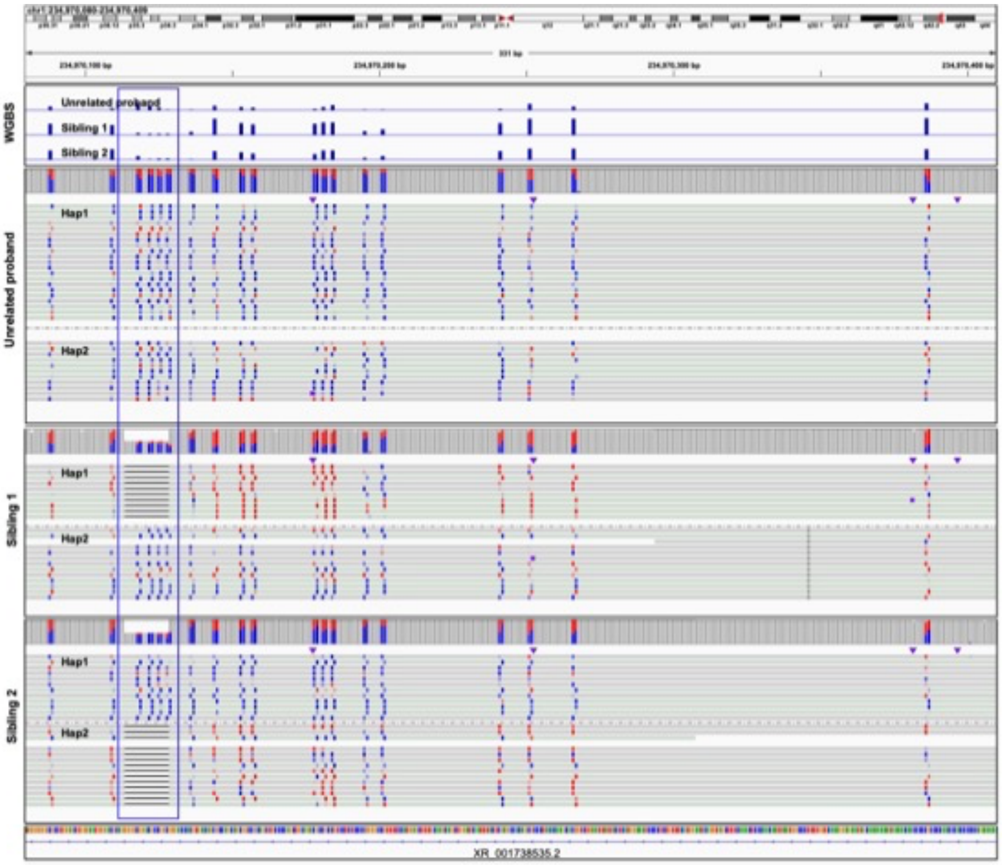

**B**

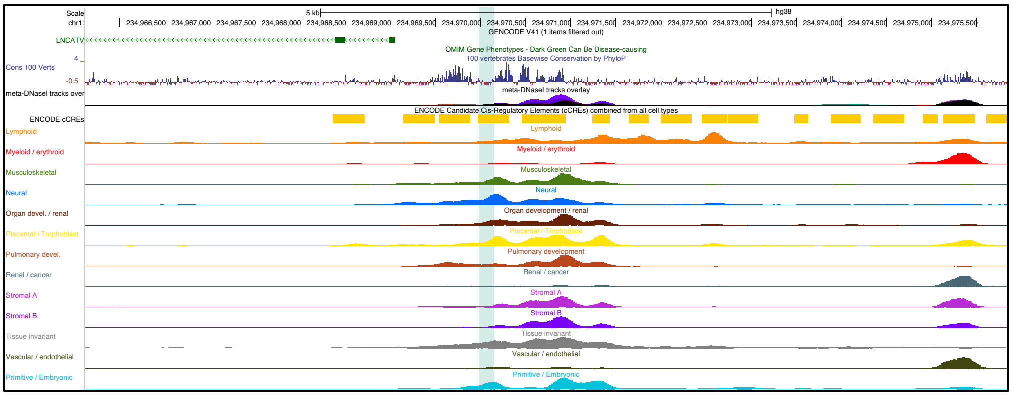

**Supplementary Figure 19. A.** Genomics view of the *FES* locus showing differential expression of T and G alleles (red box; Fisher's exact test P=0.008). **B.** Genomics view of the *NUP153* locus showing differential expression of A and G alleles (red box; Fisher's exact test P=0.005). Tracks depict full length cDNA (IsoSeq) sequence reads generated from proband-specific blood-derived iPSC lines.

**A**

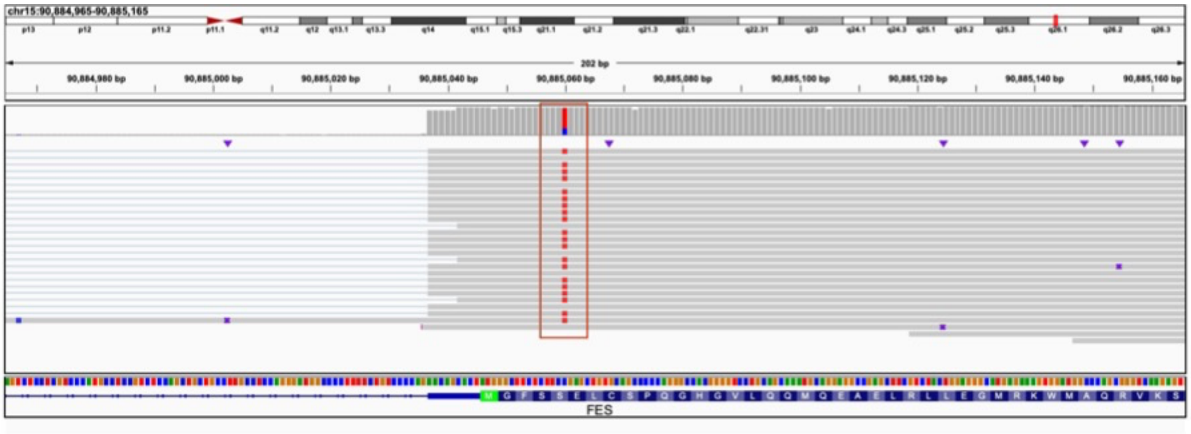

**B**

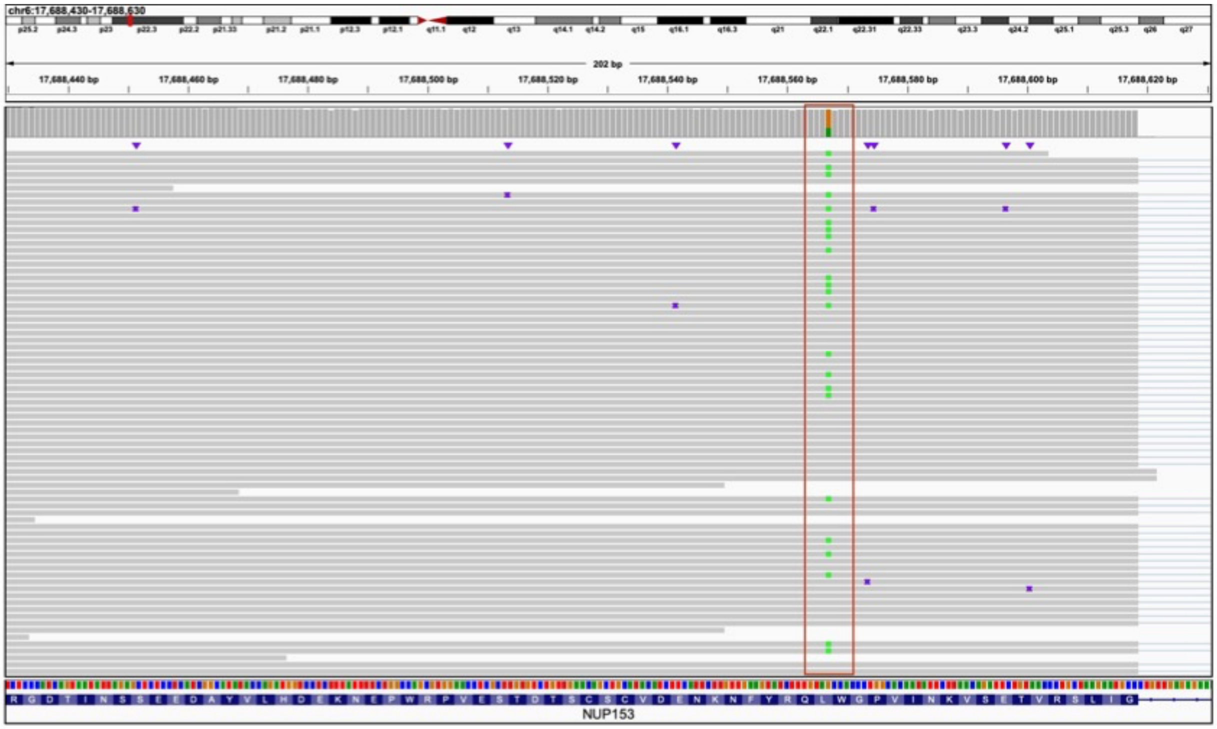

**Supplementary Figure 20.** Bulk tissue gene expression for *GNAO1* (ENSG00000087258.14) from GTEx Analysis Release V8 (dbGaP Accession phs000424.v8.p2). Expression values (y axis) are shown in TPM (transcripts per million) calculated from a gene model with isoforms collapsed to a single gene. Box plots are shown as median and 25th and 75th percentiles; points are displayed as outliers if they are above or below 1.5 times the interquartile range.

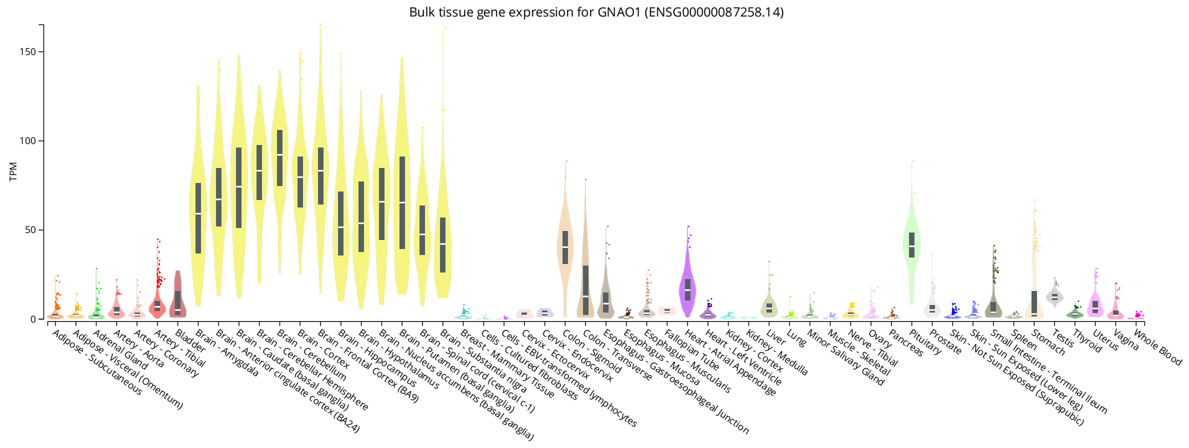
